# Prevalent comorbidities among young and underprivileged: Death portrait of COVID-19 among 235 555 hospitalized patients in Brazil

**DOI:** 10.1101/2021.01.22.21250346

**Authors:** Eduardo Pietre, Gabriel P Amorim, Maria F F Bittencourt, Marcelo Ribeiro-Alves, Mariana Acquarone

## Abstract

COVID-19 has been alarmingly spreading worldwide, with Brazil ranking third in total number of cases and second in deaths. Being a continental country, which comprises many ethnic groups and an engrained social inequality, the pandemic evidenced this heterogeneous discrepancy. We aimed to estimate the impact of associated risk factors, isolated or combined, on COVID-19 severeness, detecting specific epidemiological profiles for multiple age ranges in hospitalized Brazilians.

**Methods:** In this large retrospective cohort study, we used open-access data from the Ministry of Health of Brazil with COVID-19 confirmed hospitalized patients annotated in SRAG system between February and August 2020, a total of 235555 entries. The association of COVID-19 death with socio-demographic and clinical characteristics was analysed and presented as odds ratios adjusted by confounding co-variables. We also presented marginal mean aOR values for high-order interactions either by or not another fixed level or condition. We kept all other variables in the multivariate logistic models in their mean values or equal proportions.

**Findings:** Younger individuals with one or more comorbidities had an adjusted odds ratio up to four-fold compared to those without it, in the same age interval. Younger diabetic patients either self-declared as brown ethnicity (aOR 5·58, 95% CI 4·97-6·25; p<0·0001) or with some other associated comorbidities, mainly chronic hematologic disease (21·09, 13·64-32·6; p<0·0001) and obesity (aOR 21·7, 95% CI not calculated; p<0·0001), resulted in outstanding death risk. Age over 60, particularly over 90 (28·91, 24·5-34·11; p<0.001), usage of invasive ventilatory support (16·23, 14·05-18·75; p<0·001), admission to intensive care units (3·14, 2·82-3·48; p<0·001), multiple respiratory symptoms (3·24, 2·79-3·75; p<0·0001), black ethnicity (1·78, 1·52-2·07; p<0·05), and diagnosis previous to hospitalization (1·32, 1·19-1·47; p<0·05) were associated with higher death odds. As protective factors, with roughly one third less death risk, we found hospitalization duration of (4, 7] days and illness onset to hospitalization over 6 days.

**Interpretation:** We found evidence for increased COVID-19 risk in two distinct groups: younger patients with prevalent comorbidities, especially in brown ethnicity, and patients with black ethnicity. We speculate that the pro-inflammatory synergism of COVID-19 and comorbidities, promoting an overproduction of cytokines, is partially the cause of higher mortality in this young group. Brazilian black and brown are underprivileged populations, with structural social inequality, limited healthcare access and, thus, remarkable disease vulnerability. Our study supplies essential data to patient stratification upon admission, optimizing hospital management, and to guide public policy determinations, including group prioritization for COVID-19 vaccination in Brazil.

**Funding:** None.

**Research in context**

Evidence before this study
COVID-19 is still very active, having spread to over two hundred countries and caused more than one million deaths worldwide. Its current situation requires large-scale studies to assess the impact of preexisting comorbidities, symptoms, and socioeconomic issues regarding mortality rate, especially where lack of control is evident. We searched PubMed, Google Scholar, medRxiv, and bioRxiv on Dec 12, 2020, for studies published in English or Brazilian Portuguese, estimating the impact of several risk factors in COVID-19 prognosis. We used the search terms “Brazil” or “risk factors” or “ethnicity” or “cohort” or “diabetes mellitus” or “mortality” or “symptoms” or “comorbidities”, and related synonyms, combined with “SARS-CoV-2” or “COVID-19”. Many pre-existing conditions have shown to directly impact patient prognosis, out of which cancer, chronic kidney disease, chronic obstructive pulmonary disease, cardiovascular disease, obesity, and diabetes, among others, are well established in SARS-CoV-2 infection severeness. Some studies reported an increased death risk for non-white Brazilians, but no large scale cohort analyzing the impact of one or more associated risk factors in younger Brazilians patients were found.

Added value of this study
We found that the impact of having one or two or more risk factors on mortality are progressively higher in ages (60, 80], (40, 60], (20, 40], and (0, 20], compared with people of the same age interval without comorbidities. We also found that young brown individuals with diabetes, as well as black ethnicity on its own, are population subgroups at remarkably higher risk for severe COVID-19 in Brazil. Furthermore, advanced age, usage of ventilatory support, admission to intensive care units, multiple respiratory symptoms, and diagnosis previous to hospitalization were associated with higher death odds. As protective factors, we found hospitalization duration of (4, 7] days and illness onset to hospitalization over 6 days.

Implications of all the available evidence
We identified multiple epidemiological profiles associated with death risk in different age ranges in Brazilian COVID-19 hospitalized patients. These findings unveil that a large part of Brazilian working-age population is at a higher risk for SARS-CoV-2 death, a neglected situation that is further exacerbating inequalities, leading to a striking sociodemographic and economic impact. We hope that our analysis aids patient risk stratification, hospital management optimization, and public policy determination, including prioritization for COVID-19 vaccination in Brazil.

## INTRODUCTION

Coronavirus disease (COVID-19), the recently discovered virus named Severe Acute Respiratory Syndrome Coronavirus 2 (SARS-CoV-2), was first reported in December 2019, in Wuhan, China,^1^ and on February 25^th^, 2020, in Brazil.^2^ Due to its high transmission rate, the World Health Organization (WHO) declared, in January 2020, COVID-19 as a Public Health Emergency epidemic, and considered it a pandemic on March 11^th^, 2020.^3,4^

As of October 16^th^, 2020, more than 38619674 cases and 1093522 deaths caused by SARS-CoV-2 have been reported worldwide; 7833851 cases and 215199 deaths reported in the USA and 5140863 cases and 151747 deaths reported in Brazil, the first and third biggest epicenters, respectively.^5^ Although Brazil population accounts for only 2·75% of the world population,^6^ 13·31% of reported cases and 13·87% of reported deaths are from this country,^5^ showing a remarkable lack of transmission control effectiveness.

COVID-19 has heterogeneous clinical manifestations, ranging from mild nonspecific flu-like symptoms to severe pneumonia with multiple organ failure and death. Most common symptoms may include fever, cough, fatigue, dyspnoea, myalgia, and sputum production.^7^ Headache, gastrointestinal symptoms, anosmia, and dysgeusia can also occur.^8,9^ Cancer, chronic kidney disease, chronic obstructive pulmonary disease, cardiovascular disease, obesity, sickle cell disease, smoking, organ transplantation, and type 2 diabetes mellitus are consistently considered risk factors for COVID-19 severity.^10^

Diabetes mellitus (DM) is a global public health issue,^11^ Brazil being the fifth country in the number of affected people.^11^ DM is an aggravating factor among other respiratory coronavirus infections as well,^12,13^ and, similarly to other COVID-19 severity risk factors, leads to a chronic proinflammatory state with an attenuation of the immunity response with vascular impairment. The cytokine-driven response is highly correlated with cytokine storm, and the unbalanced immune system becomes a vital promoter of worsening of the patient’s condition.^14^

Based on data from the Health Ministry of Brazil, we aimed to estimate the impact of social demographic features, comorbidities, and clinical conditions, resulting in the identification of an epidemiological profile of COVID-19 hospitalization in Brazil. We found younger individuals with one or more comorbidities, younger diabetic self-declared as brown ethnicity, and black ethnicity patients at outstanding aggravating death risk.

## METHODS

### STUDY DESIGN AND PATIENTS

In this retrospective cohort study, an official open-source (creative commons licensed) dataset called “Severe Acute Respiratory Syndrome Database”, kept and updated by the Ministry of Health of Brazil,^15^ has been used, and its data released on August 12^th^, 2020, 19:05 (UTC-03:00), have been analysed. Only patients with positive laboratory samples (polymerase chain reaction or serological) or clinical (symptoms, imaging, or epidemiological link) COVID-19 diagnosis, hospitalized and dismissed from February 20^th^ to August 9^th^ were included in the analyses.^16^ Among 575935 records, 282495 were discarded from analyses, due to presenting a confirmed etiologic agent other than SARS-CoV-2 or being waiting for conclusive results. Furthermore, entries with sex marked as ignored (83 records), without a disclosed clinical evolution (56099 records), with age equal to 0 (4 records) or age under one year (1699 records) were also discarded, resulting in the 235555 records used in the analyses.

Records that had the “other risk factors” (in this study referred to as “unlisted risk factors”) field marked as true and text description that included “HAS”, “H.A.S.”, “HIPERTENSO”, “HIPERTENSAO” (when not “HIPERTENSAO PULMONAR”), “HIPERTENSA”, or “HA” were assigned to a new chronic high blood pressure variable (29017 records). Entries whose description included “OBESIDADE”, “OBESO”, and “OBESA” were assigned to obesity (4854 records) as a standard term. In cases where this was the only description for unlisted risk factors, the variable was changed to false (25004 records).

### DATA COLLECTION

COVID-19 monitoring in Brazil is made possible due to the repurposing of two sentinel surveillance systems created by the Ministry of Health: influenza-like illness (denominated flu-like syndrome) and severe acute respiratory syndrome tracking (here abbreviated as SRAG). The compulsory notification occurs on the online platform “e-SUS Notifica”, nationally, made by health professionals from public and private institutions. All influenza and influenza-like illnesses are monitored, annotated, and compiled in the SIVEP-GRIPE database,^17^ with government restricted access as of August 12, 2020. Cases of hospitalizations or deaths due to acute respiratory syndrome severeness are also annotated in the open-access SRAG database.^17^ According to the Ministry of Health of Brazil, the classification of SRAG demands the patient fulfills the flu-like syndrome definition and manifests dyspnea, respiratory distress, chest pain, oxygen saturation less than 95%, or cyanosis.^17^ In children, particularly, intercostal retraction, dehydration, and inappetence are also considered.^17^

### SEVERE ACUTE RESPIRATORY SYNDROME FORM

The SRAG form comprises 154 variables, including extensive sociodemographic and clinical information, some limited to the government. The form has mandatory fields for annotation in the Information System for Notifiable Diseases (SINAN). An extra field is also available for complementary observations. Out of those, we selected the following for the analyses: annotation date, symptoms offset date, case disclosure date, hospitalization date, sex, age, ethnicity, schooling level, pregnancy, fever, cough, sore throat, dyspnea, respiratory distress, saturation level, diarrhea, vomiting, presence of risk factors, postpartum period, chronic cardiovascular disease, chronic hematologic disease, down syndrome, chronic hepatic disease, asthma, diabetes mellitus, chronic neurological disease, immunodeficiency or immunosuppression, chronic renal disease, obesity, obesity body mass index, unlisted risk factors, ventilatory support, chest x-ray, computerized tomography, intensive care unit, previously known diagnosis, final diagnosis, and patient outcome.

### STATISTICAL ANALYSIS

For the description of the patients who died or not due to COVID-19 and its consequences up to the time of the study, according to their socio-demographic and clinical characteristics, for continuous numerical variables, we used density plots, medians, and interquartile intervals, and performed non-parametric Mann-Whitney tests comparing central tendencies; for nominal/categorical variables, we used bar plots, calculated absolute and relative frequencies, and tested independence with Fisher’s exact tests.

In determining factors associated with death risk and protection, we used fixed-effect generalized linear parametric models with a logistic link function (Binomial family). The measure of association was presented as a function of odds ratios either adjusted by co-variables of confounding (aOR) or not (OR). P-values were corrected by the number of comparisons with the reference level by the Holm-Sidak method. We kept all other variables in the multivariate logistic models in their mean values or equal proportions to estimate marginal mean aOR values and its 95% confidence intervals for high-order interactions either by or not another fixed level or condition. Again, p-values were corrected by the number of comparisons with the reference level by the Holm-Sidak method.

All statistical analyses were performed in the software R v.3.6.3 and the library ‘emmeans’ and its dependencies.

## RESULTS

Out of the 181964 patients, 56·3% were males and 43·7% females (Table 1). Of 96567 deaths, 58·23% were among males and 41·77% among females, resulting in 41% mortality (Table 1). The median age was 61 (IQR=28), the mean age 59·44, and the standard deviation 18·53 (Table 1). The ethnic analysis shows that 31% were self-declared as white, 4·6% black, 1·1% yellow, 32·4% brown, 0·3% red (indigenous), and 30·6% blank or ignored (Table 1). Schooling level demonstrated 2·5% were illiterate, 8·8% completed Elementary school only partially, 6·3% completed Elementary school, 11·2% completed High school, and 5·3% graduated from College. Over half of schooling level entries were missing or marked as ignored (Table 1). Pregnancy stage analysis revealed 31·4% of patients were not pregnant, 0·1% were in the first trimester of gestational age, 0·2% were in the second trimester, 0·6% in the third trimester, and 0·1% were pregnant but with no disclosed gestational age (Table 1).

**Table 1:** Sociodemographic findings of patients on admission.

| Feature | Category | Overall | Cure | Death | P-value |
| --- | --- | --- | --- | --- | --- |
| Sex |  |  |  |  | < 0.001 |
|  | Female | 103014 (43.7%) | 62680 (26.6%) | 40334 (17.1%) |  |
|  | Male | 132541 (56.3%) | 76308 (32.4%) | 56233 (23.9%) |  |
| Age |  |  |  |  | < 0.001 |
|  |  | 61 (IQR=28) | 53 (IQR=26) | 70 (IQR=21) |  |
| Age Range |  |  |  |  | < 0.001 |
|  | (0, 10] | 1804 (0.8%) | 1670 (0.7%) | 134 (0.1%) |  |
|  | (10, 20] | 2493 (1.1%) | 2181 (0.9%) | 312 (0.1%) |  |
|  | (20, 30] | 10942 (4.6%) | 9784 (4.2%) | 1158 (0.5%) |  |
|  | (30, 40] | 25329 (10.8%) | 21726 (9.2%) | 3603 (1.5%) |  |
|  | (40, 50] | 34067 (14.5%) | 26739 (11.4%) | 7328 (3.1%) |  |
|  | (50, 60] | 42101 (17.9%) | 28348 (12%) | 13753 (5.8%) |  |
|  | (60, 70] | 46675 (19.8%) | 23951 (10.2%) | 22724 (9.6%) |  |
|  | (70, 80] | 40130 (17%) | 15653 (6.6%) | 24477 (10.4%) |  |
|  | (80, 90] | 25646 (10.9%) | 7547 (3.2%) | 18099 (7.7%) |  |
|  | (90, 100] | 6090 (2.6%) | 1349 (0.6%) | 4741 (2%) |  |
|  | (100, 110] | 273 (0.1%) | 39 (0%) | 234 (0.1%) |  |
|  | > 110 | 5 (0%) | 1 (0%) | 4 (0%) |  |
| Ethnicity |  |  |  |  | < 0.001 |
|  | White | 72913 (31%) | 44949 (19.1%) | 27964 (11.9%) |  |
|  | Black | 10870 (4.6%) | 5863 (2.5%) | 5007 (2.1%) |  |
|  | Yellow | 2539 (1.1%) | 1453 (0.6%) | 1086 (0.5%) |  |
|  | Brown | 76311 (32.4%) | 41890 (17.8%) | 34421 (14.6%) |  |
|  | Red | 734 (0.3%) | 367 (0.2%) | 367 (0.2%) |  |
|  | Unknown | 72188 (30.6%) | 44466 (18.9%) | 27722 (11.8%) |  |
| Schooling level |  |  |  |  | < 0.001 |
|  | 0 years | 5976 (2.5%) | 2112 (0.9%) | 3864 (1.6%) |  |
|  | 5 years | 20795 (8.8%) | 10165 (4.3%) | 10630 (4.5%) |  |
|  | 9 years | 14823 (6.3%) | 8508 (3.6%) | 6315 (2.7%) |  |
|  | 12 years | 26320 (11.2%) | 18602 (7.9%) | 7718 (3.3%) |  |
|  | Graduated | 12468 (5.3%) | 9695 (4.1%) | 2773 (1.2%) |  |
|  | N.A. | 608 (0.3%) | 561 (0.2%) | 47 (0%) |  |
|  | Unknown | 154565 (65.6%) | 89345 (37.9%) | 65220 (27.7%) |  |
| Pregnancy |  |  |  |  | < 0.001 |
|  | No | 74029 (31.4%) | 45485 (19.3%) | 28544 (12.1%) |  |
|  | 1st Trimester | 176 (0.1%) | 169 (0.1%) | 7 (0%) |  |
|  | 2nd Trimester | 490 (0.2%) | 439 (0.2%) | 51 (0%) |  |
|  | 3rd Trimester | 1335 (0.6%) | 1248 (0.5%) | 87 (0%) |  |
|  | Unknown Trimester | 125 (0.1%) | 113 (0%) | 12 (0%) |  |
|  | N.A. | 147806 (62.8%) | 83851 (35.6%) | 63955 (27.2%) |  |
|  | Unknown | 11575 (4.9%) | 7668 (3.3%) | 3907 (1.7%) |  |

Age represented as median (IQR) and proportions represented by n (%). Schooling level expressed as the number of years of study. P-values were calculated either by Mann-Whitney or Fisher's exact tests. N.A.=Not Applicable.

Only 22·9% of patients were diagnosed and notified as COVID-19 before requiring hospitalization, whereas 48·1% were assigned as COVID-19 positive just after, and this information was marked as ignored in 29% of cases (Table 2). The median hospitalization duration was 7 days (IQR=9), mean 10·11 days, and standard deviation 10·26 (Table 2). The median time from illness onset to hospitalization was 6 days (IQR=6), and the median time from illness onset to cure or death was 14 days (IQR=11) (Table 2). The median notification delay was 0 days (IQR=2) (Table 2). Half the patients did not use intensive care unit, while 30·2% used it, and 19·6% had this information blank or marked as ignored (Table 2). Ventilatory support was not used by 24·5%, invasive ventilatory support by 18·5%, and non-invasive ventilatory support by 37·5% of patients (Table 2).

**Table 2:** Clinical and radiologic finding of patients.

| Feature | Category | Overall | Cure | Death | P-value |
| --- | --- | --- | --- | --- | --- |
| Fever |  |  |  |  | < 0.001 |
|  | No | 48316 (20.5%) | 27543 (11.7%) | 20773 (8.8%) |  |
|  | Yes | 158847 (67.4%) | 98796 (41.9%) | 60051 (25.5%) |  |
|  | Unknown | 28392 (12.1%) | 12649 (5.4%) | 15743 (6.7%) |  |
| Cough |  |  |  |  | < 0.001 |
|  | No | 38100 (16.2%) | 21042 (8.9%) | 17058 (7.2%) |  |
|  | Yes | 171405 (72.8%) | 106745 (45.3%) | 64660 (27.5%) |  |
|  | Unknown | 26050 (11.1%) | 11201 (4.8%) | 14849 (6.3%) |  |
| Sore throat |  |  |  |  | < 0.001 |
|  | No | 124521 (52.9%) | 74822 (31.8%) | 49699 (21.1%) |  |
|  | Yes | 46117 (19.6%) | 31003 (13.2%) | 15114 (6.4%) |  |
|  | Unknown | 64917 (27.6%) | 33163 (14.1%) | 31754 (13.5%) |  |
| Dyspnea |  |  |  |  | < 0.001 |
|  | No | 44237 (18.8%) | 32928 (14%) | 11309 (4.8%) |  |
|  | Yes | 162640 (69%) | 89491 (38%) | 73149 (31.1%) |  |
|  | Unknown | 28678 (12.2%) | 16569 (7%) | 12109 (5.1%) |  |
| Respiratory distress |  |  |  |  | < 0.001 |
|  | No | 60879 (25.8%) | 43642 (18.5%) | 17237 (7.3%) |  |
|  | Yes | 129968 (55.2%) | 69439 (29.5%) | 60529 (25.7%) |  |
|  | Unknown | 44708 (19%) | 25907 (11%) | 18801 (8%) |  |
| Oxygen saturation < 95% |  |  |  |  | < 0.001 |
|  | No | 62311 (26.5%) | 46972 (19.9%) | 15339 (6.5%) |  |
|  | Yes | 128895 (54.7%) | 65884 (28%) | 63011 (26.8%) |  |
|  | Unknown | 44349 (18.8%) | 26132 (11.1%) | 18217 (7.7%) |  |
| Diarrhea |  |  |  |  | < 0.001 |
|  | No | 135235 (57.4%) | 81912 (34.8%) | 53323 (22.6%) |  |
|  | Yes | 31053 (13.2%) | 20931 (8.9%) | 10122 (4.3%) |  |
|  | Unknown | 69267 (29.4%) | 36145 (15.3%) | 33122 (14.1%) |  |
| Vomiting |  |  |  |  | < 0.001 |
|  | No | 144483 (61.3%) | 88567 (37.6%) | 55916 (23.7%) |  |
|  | Yes | 17752 (7.5%) | 11477 (4.9%) | 6275 (2.7%) |  |
|  | Unknown | 73320 (31.1%) | 38944 (16.5%) | 34376 (14.6%) |  |
| Risk Factor |  |  |  |  | < 0.001 |
|  | No | 89699 (38.1%) | 63309 (26.9%) | 26390 (11.2%) |  |
|  | Yes | 145856 (61.9%) | 75679 (32.1%) | 70177 (29.8%) |  |
| Postpartum period* |  |  |  |  | < 0.001 |
|  | No | 212159 (90.1%) | 126458 (53.7%) | 85701 (36.4%) |  |
|  | Yes | 621 (0.3%) | 484 (0.2%) | 137 (0.1%) |  |
|  | Unknown | 22775 (9.7%) | 12046 (5.1%) | 10729 (4.6%) |  |
| Chronic Cardiovascular Disease |  |  |  |  | < 0.001 |
|  | No | 130153 (55.3%) | 86280 (36.6%) | 43873 (18.6%) |  |
|  | Yes | 75916 (32.2%) | 37500 (15.9%) | 38416 (16.3%) |  |
|  | Unknown | 29486 (12.5%) | 15208 (6.5%) | 14278 (6.1%) |  |
| Chronic Hematologic Disease |  |  |  |  | < 0.001 |
|  | No | 172689 (73.3%) | 107620 (45.7%) | 65069 (27.6%) |  |
|  | Yes | 1903 (0.8%) | 911 (0.4%) | 992 (0.4%) |  |
|  | Unknown | 60963 (25.9%) | 30457 (12.9%) | 30506 (13%) |  |
| Down Syndrome |  |  |  |  | < 0.001 |
|  | No | 173851 (73.8%) | 108151 (45.9%) | 65700 (27.9%) |  |
|  | Yes | 571 (0.2%) | 271 (0.1%) | 300 (0.1%) |  |
|  | Unknown | 61133 (26%) | 30566 (13%) | 30567 (13%) |  |
| Chronic Hepatic Disease |  |  |  |  | < 0.001 |
|  | No | 172113 (73.1%) | 107413 (45.6%) | 64700 (27.5%) |  |
|  | Yes | 2087 (0.9%) | 841 (0.4%) | 1246 (0.5%) |  |
|  | Unknown | 61355 (26%) | 30734 (13%) | 30621 (13%) |  |
| Asthma |  |  |  |  | < 0.001 |
|  | No | 169828 (72.1%) | 105508 (44.8%) | 64320 (27.3%) |  |
|  | Yes | 5964 (2.5%) | 3980 (1.7%) | 1984 (0.8%) |  |
|  | Unknown | 59763 (25.4%) | 29500 (12.5%) | 30263 (12.8%) |  |
| Diabetes Mellitus |  |  |  |  | < 0.001 |
|  | No | 139666 (59.3%) | 91270 (38.7%) | 48396 (20.5%) |  |
|  | Yes | 58764 (24.9%) | 28364 (12%) | 30400 (12.9%) |  |
|  | Unknown | 37125 (15.8%) | 19354 (8.2%) | 17771 (7.5%) |  |
| Chronic Neurological Disease |  |  |  |  | < 0.001 |
|  | No | 168243 (71.4%) | 106170 (45.1%) | 62073 (26.4%) |  |
|  | Yes | 9071 (3.9%) | 3326 (1.4%) | 5745 (2.4%) |  |
|  | Unknown | 58241 (24.7%) | 29492 (12.5%) | 28749 (12.2%) |  |
| Chronic Pneumopathy |  |  |  |  | < 0.001 |
|  | No | 168305 (71.5%) | 106011 (45%) | 62294 (26.4%) |  |
|  | Yes | 8534 (3.6%) | 3314 (1.4%) | 5220 (2.2%) |  |
|  | Unknown | 58716 (24.9%) | 29663 (12.6%) | 29053 (12.3%) |  |
| Immunodeficiency /<br>Immunosuppression |  |  |  |  | < 0.001 |
|  | No | 169112 (71.8%) | 105999 (45%) | 63113 (26.8%) |  |
|  | Yes | 6460 (2.7%) | 2965 (1.3%) | 3495 (1.5%) |  |
|  | Unknown | 59983 (25.5%) | 30024 (12.7%) | 29959 (12.7%) |  |
| Chronic Renal Disease |  |  |  |  | < 0.001 |
|  | No | 167141 (71%) | 105644 (44.8%) | 61497 (26.1%) |  |
|  | Yes | 9820 (4.2%) | 3522 (1.5%) | 6298 (2.7%) |  |
|  | Unknown | 58594 (24.9%) | 29822 (12.7%) | 28772 (12.2%) |  |
| Obesity |  |  |  |  | < 0.001 |
|  | No | 163198 (69.3%) | 102374 (43.5%) | 60824 (25.8%) |  |
|  | Yes | 13940 (5.9%) | 7703 (3.3%) | 6237 (2.6%) |  |
|  | Unknown | 58417 (24.8%) | 28911 (12.3%) | 29506 (12.5%) |  |
| High Blood Pressure |  |  |  |  | < 0.001 |
|  | No | 89699 (38.1%) | 63309 (26.9%) | 26390 (11.2%) |  |
|  | Yes | 29017 (12.3%) | 14661 (6.2%) | 14356 (6.1%) |  |
|  | Unknown | 116839 (49.6%) | 61018 (25.9%) | 55821 (23.7%) |  |
| Unlisted risk factors |  |  |  |  | < 0.001 |
|  | No | 156916 (66.6%) | 99739 (42.3%) | 57177 (24.3%) |  |
|  | Yes | 37370 (15.9%) | 18154 (7.7%) | 19216 (8.2%) |  |
|  | Unknown | 41269 (17.5%) | 21095 (9%) | 20174 (8.6%) |  |
| Number of risk factors |  |  |  |  | < 0.001 |
|  | 0 | 90282 (38.3%) | 63737 (27.1%) | 26545 (11.3%) |  |
|  | 1 | 68017 (28.9%) | 39438 (16.7%) | 28579 (12.1%) |  |
|  | 2 | 49711 (21.1%) | 24416 (10.4%) | 25295 (10.7%) |  |
|  | 3 | 19933 (8.5%) | 8635 (3.7%) | 11298 (4.8%) |  |
|  | 4 | 5805 (2.5%) | 2194 (0.9%) | 3611 (1.5%) |  |
|  | 5 | 1433 (0.6%) | 451 (0.2%) | 982 (0.4%) |  |
|  | 6 | 293 (0.1%) | 83 (0%) | 210 (0.1%) |  |
|  | 7 | 44 (0%) | 11 (0%) | 33 (0%) |  |
|  | 8 | 14 (0%) | 9 (0%) | 5 (0%) |  |
|  | 9 | 8 (0%) | 5 (0%) | 3 (0%) |  |
|  | 10 | 5 (0%) | 4 (0%) | 1 (0%) |  |
|  | 11 | 6 (0%) | 3 (0%) | 3 (0%) |  |
|  | 12 | 3 (0%) | 1 (0%) | 2 (0%) |  |
|  | 13 | 1 (0%) | 1 (0%) | 0 (0%) |  |
| Ventilatory Support |  |  |  |  | < 0.001 |
|  | No | 57750 (24.5%) | 47265 (20.1%) | 10485 (4.5%) |  |
|  | Yes, invasive | 43647 (18.5%) | 7248 (3.1%) | 36399 (15.5%) |  |
|  | Yes, non-invasive | 88419 (37.5%) | 60862 (25.8%) | 27557 (11.7%) |  |
|  | Unknown | 45739 (19.4%) | 23613 (10%) | 22126 (9.4%) |  |
| X-ray |  |  |  |  | < 0.001 |
|  | Normal | 5175 (2.2%) | 3692 (1.6%) | 1483 (0.6%) |  |
|  | Interstitial infiltrate | 39480 (16.8%) | 23453 (10%) | 16027 (6.8%) |  |
|  | Consolidation | 5716 (2.4%) | 3173 (1.3%) | 2543 (1.1%) |  |
|  | Mixed | 6904 (2.9%) | 3815 (1.6%) | 3089 (1.3%) |  |
|  | Other | 37728 (16%) | 24473 (10.4%) | 13255 (5.6%) |  |
|  | Unrealized | 40684 (17.3%) | 25804 (11%) | 14880 (6.3%) |  |
|  | Unknown | 99868 (42.4%) | 54578 (23.2%) | 45290 (19.2%) |  |
| Computerized tomography |  |  |  |  | < 0.001 |
|  | No | 62 (0.5%) | 48 (0%) | 14 (0%) |  |
|  | COVID-19 typical | 6275 (48.7%) | 3672 (1.6%) | 2603 (1.1%) |  |
|  | COVID-19 undetermined | 283 (2.2%) | 136 (0.1%) | 147 (0.1%) |  |
|  | COVID-19 atypical | 136 (1.1%) | 63 (0%) | 73 (0%) |  |
|  | Other | 651 (5.1%) | 399 (0.2%) | 252 (0.1%) |  |
|  | Unrealized | 3577 (27.8%) | 2010 (0.9%) | 1567 (0.7%) |  |
|  | Unknown | 1893 (14.7%) | 995 (0.4%) | 898 (0.4%) |  |
| Intensive Care Unit |  |  |  |  | < 0.001 |
|  | No | 118289 (50.2%) | 87797 (37.3%) | 30492 (12.9%) |  |
|  | Yes | 71190 (30.2%) | 26166 (11.1%) | 45024 (19.1%) |  |
|  | Unknown | 46076 (19.6%) | 25025 (10.6%) | 21051 (8.9%) |  |
| Know grippal case† |  |  |  |  | < 0.001 |
|  | No | 113275 (48.1%) | 71856 (30.5%) | 41419 (17.6%) |  |
|  | Yes | 54041 (22.9%) | 29837 (12.7%) | 24204 (10.3%) |  |
|  | Unknown | 68239 (29%) | 37295 (15.8%) | 30944 (13.1%) |  |
| Illness onset to hospitalization |  |  |  |  | < 0.001 |
|  | Days | 6 (IQR=6) | 6 (IQR=6) | 5 (IQR=6) |  |
| Illness onset to cure or death |  |  |  |  |  |
|  | Days | 14 (IQR=11) | 15 (IQR=10) | 13 (IQR=14) | < 0.001 |
| Hospitalization duration |  |  |  |  |  |
|  | Days | 7 (IQR=9) | 7 (IQR=8) | 8 (IQR=11) |  |
| Notification delay |  |  |  |  | < 0.001 |
|  | Days | 0 (IQR=2) | 0 (IQR=2) | 1 (IQR=3) |  |

Time duration (in days) represented as medians (IQR) and proportions represented by n (%). P-values were calculated either by Mann-Whitney or Fisher's exact tests. COVID-19=2019 novel coronavirus disease. \*Up to 45 days after giving birth. †Diagnosed and notified as COVID-19 before hospitalization.

Cough was the most reported symptom on admission, followed by dyspnea, fever, respiratory distress, oxygen saturation < 95%, sore throat, diarrhea, and vomiting (Table 2). Some risk factors were present in over half of patients. Chronic cardiovascular diseases were the most prevalent, followed by diabetes mellitus, unlisted risk factors, and high blood pressure (Table 2). Less frequent comorbidities were obesity, chronic renal disease, chronic neurological disease, chronic pneumopathy, immunodeficiency or immunosuppression, asthma, chronic hepatic disease, chronic hematologic disease, postpartum period, and down syndrome, respectively (Table 2).

Critical factors which increasingly impact outcomes, found as statistically significant by logistic analysis, were: respiratory distress, oxygen saturation < 95%, patients diagnosed with COVID-19 prior to hospitalization, dyspnea, use of non-invasive ventilatory support, black ethnicity, pre-existing chronic neurological disease, pre-existing immunodeficiency or immunosuppression, need of intensive care unit, age (60, 90], usage of invasive ventilatory support, and age greater than 90 (Table 3; Figure 1). As protective factors, illness onset prior to hospitalization greater than 6 but less than or equal 9 days, greater than 9 days, and patients with a hospitalization duration greater than 4 but less than or equal to 7 days (Table 3; Figure 1).

**Table 3:**
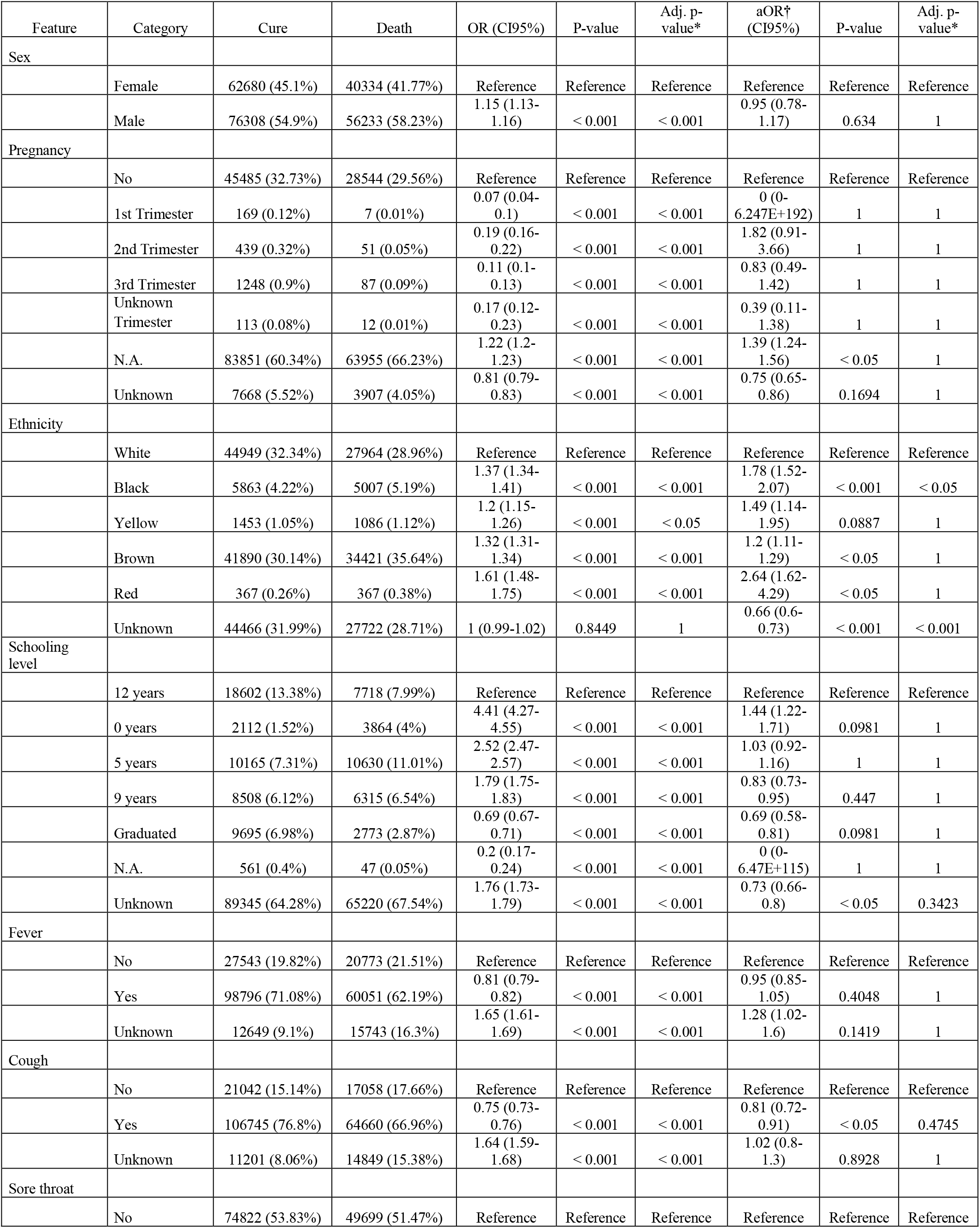
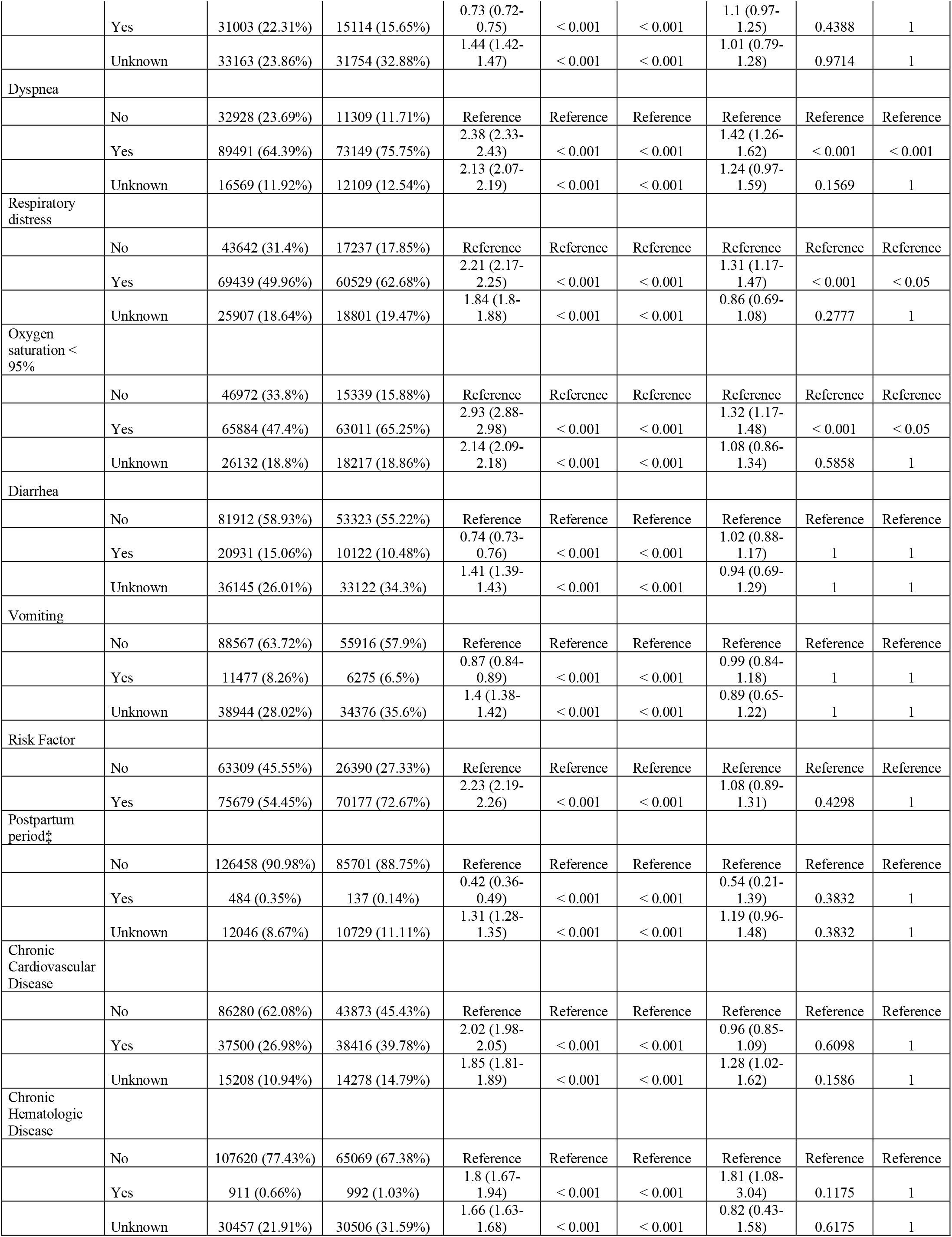
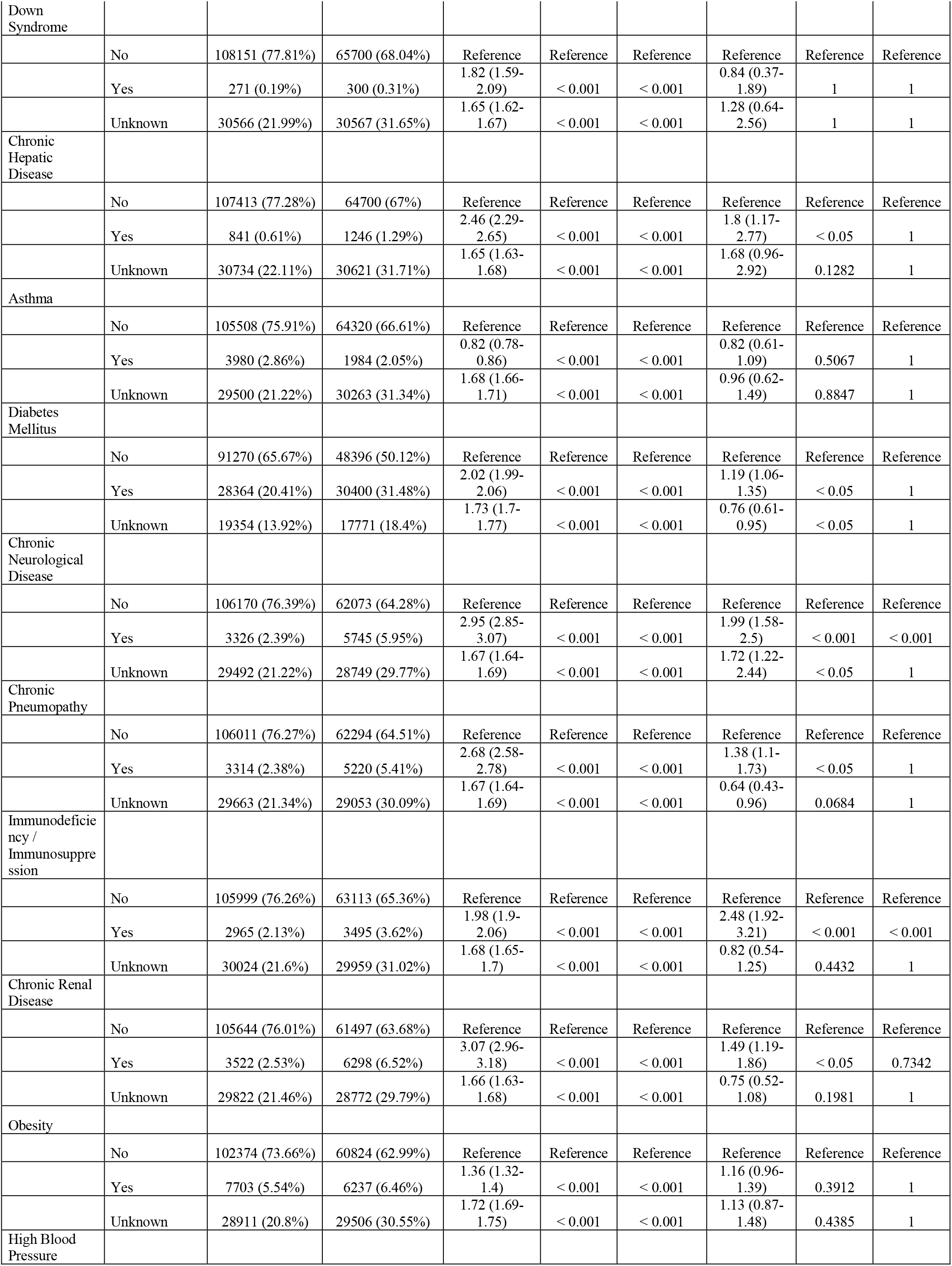
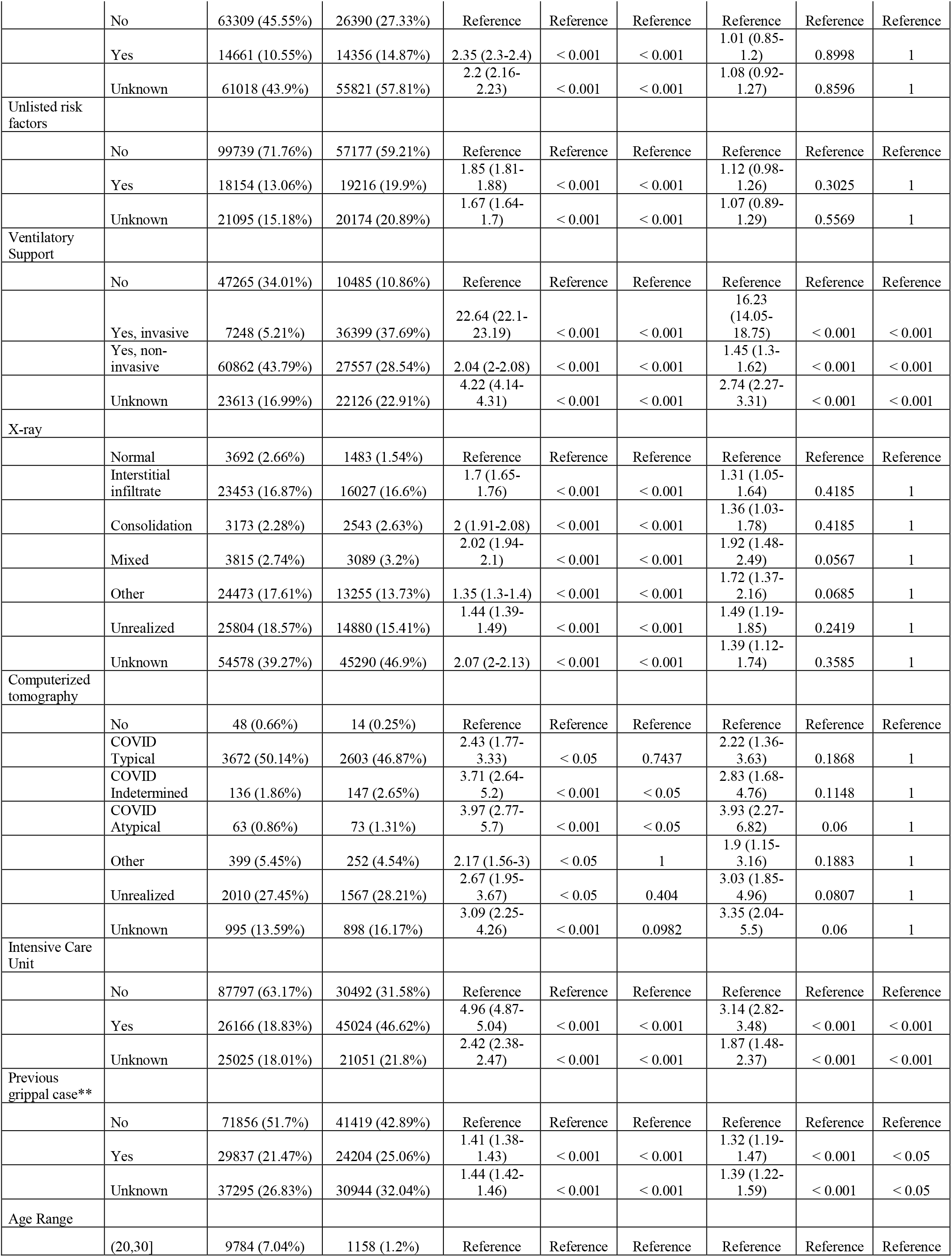
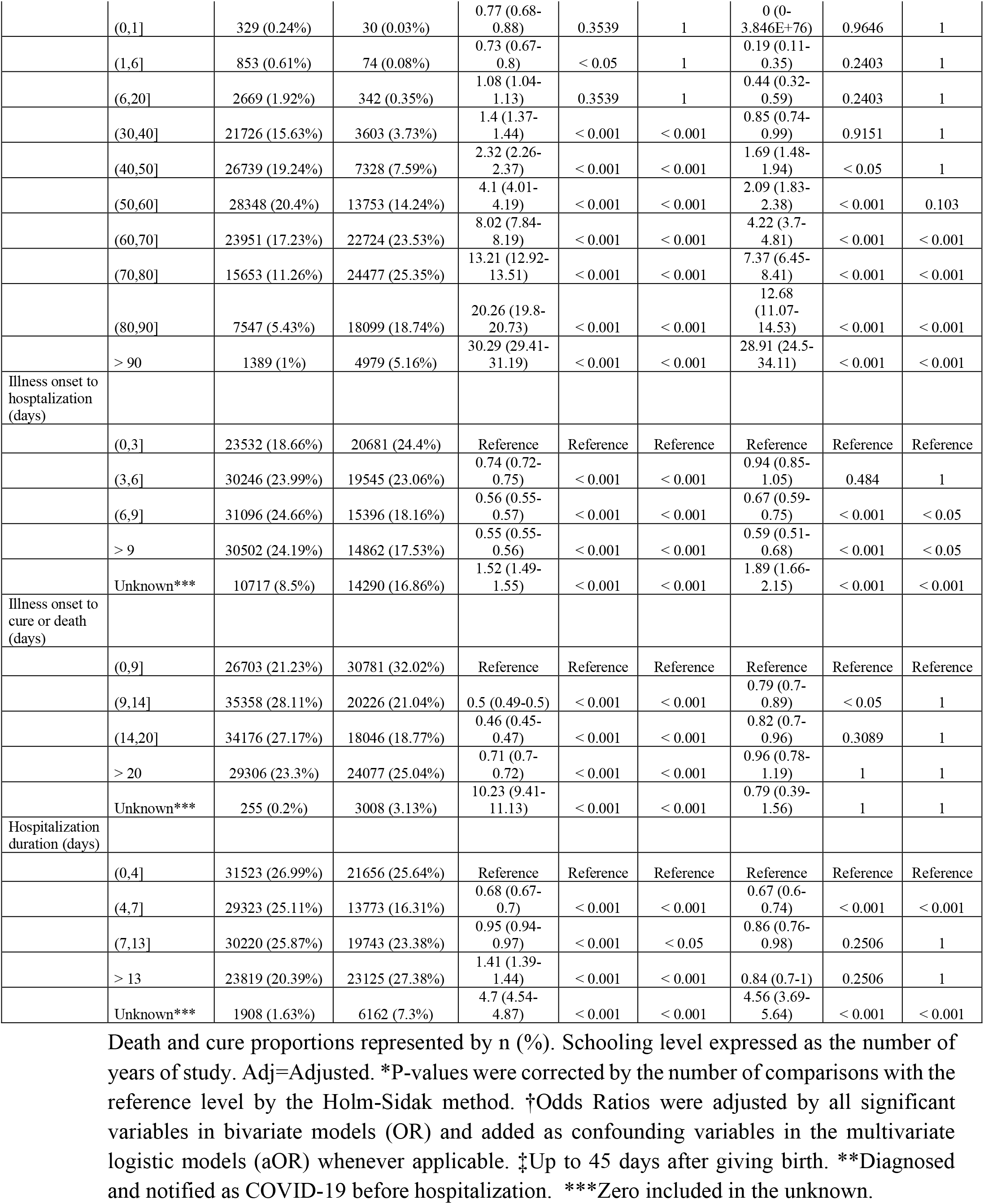
Logistic analysis for COVID-19 death.

| Feature | Category | Cure | Death | OR (CI95%) | P-value | Adj. p-value* | aOR† (CI95%) | P-value | Adj. p-value* |
| --- | --- | --- | --- | --- | --- | --- | --- | --- | --- |
| Sex |  |  |  |  |  |  |  |  |  |
|  | Female | 62680 (45.1%) | 40334 (41.77%) | Reference | Reference | Reference | Reference | Reference | Reference |
|  | Male | 76308 (54.9%) | 56233 (58.23%) | 1.15 (1.13-1.16) | < 0.001 | < 0.001 | 0.95 (0.78-1.17) | 0.634 | 1 |
| Pregnancy |  |  |  |  |  |  |  |  |  |
|  | No | 45485 (32.73%) | 28544 (29.56%) | Reference | Reference | Reference | Reference | Reference | Reference |
|  | 1st Trimester | 169 (0.12%) | 7 (0.01%) | 0.07 (0.04-0.1) | < 0.001 | < 0.001 | 0 (0-6.247E+192) | 1 | 1 |
|  | 2nd Trimester | 439 (0.32%) | 51 (0.05%) | 0.19 (0.16-0.22) | < 0.001 | < 0.001 | 1.82 (0.91-3.66) | 1 | 1 |
|  | 3rd Trimester | 1248 (0.9%) | 87 (0.09%) | 0.11 (0.1-0.13) | < 0.001 | < 0.001 | 0.83 (0.49-1.42) | 1 | 1 |
|  | Unknown Trimester | 113 (0.08%) | 12 (0.01%) | 0.17 (0.12-0.23) | < 0.001 | < 0.001 | 0.39 (0.11-1.38) | 1 | 1 |
|  | N.A. | 83851 (60.34%) | 63955 (66.23%) | 1.22 (1.2-1.23) | < 0.001 | < 0.001 | 1.39 (1.24-1.56) | < 0.05 | 1 |
|  | Unknown | 7668 (5.52%) | 3907 (4.05%) | 0.81 (0.79-0.83) | < 0.001 | < 0.001 | 0.75 (0.65-0.86) | 0.1694 | 1 |
| Ethnicity |  |  |  |  |  |  |  |  |  |
|  | White | 44949 (32.34%) | 27964 (28.96%) | Reference | Reference | Reference | Reference | Reference | Reference |
|  | Black | 5863 (4.22%) | 5007 (5.19%) | 1.37 (1.34-1.41) | < 0.001 | < 0.001 | 1.78 (1.52-2.07) | < 0.001 | < 0.05 |
|  | Yellow | 1453 (1.05%) | 1086 (1.12%) | 1.2 (1.15-1.26) | < 0.001 | < 0.05 | 1.49 (1.14-1.95) | 0.0887 | 1 |
|  | Brown | 41890 (30.14%) | 34421 (35.64%) | 1.32 (1.31-1.34) | < 0.001 | < 0.001 | 1.2 (1.11-1.29) | < 0.05 | 1 |
|  | Red | 367 (0.26%) | 367 (0.38%) | 1.61 (1.48-1.75) | < 0.001 | < 0.001 | 2.64 (1.62-4.29) | < 0.05 | 1 |
|  | Unknown | 44466 (31.99%) | 27722 (28.71%) | 1 (0.99-1.02) | 0.8449 | 1 | 0.66 (0.6-0.73) | < 0.001 | < 0.001 |
| Schooling level |  |  |  |  |  |  |  |  |  |
|  | 12 years | 18602 (13.38%) | 7718 (7.99%) | Reference | Reference | Reference | Reference | Reference | Reference |
|  | 0 years | 2112 (1.52%) | 3864 (4%) | 4.41 (4.27-4.55) | < 0.001 | < 0.001 | 1.44 (1.22-1.71) | 0.0981 | 1 |
|  | 5 years | 10165 (7.31%) | 10630 (11.01%) | 2.52 (2.47-2.57) | < 0.001 | < 0.001 | 1.03 (0.92-1.16) | 1 | 1 |
|  | 9 years | 8508 (6.12%) | 6315 (6.54%) | 1.79 (1.75-1.83) | < 0.001 | < 0.001 | 0.83 (0.73-0.95) | 0.447 | 1 |
|  | Graduated | 9695 (6.98%) | 2773 (2.87%) | 0.69 (0.67-0.71) | < 0.001 | < 0.001 | 0.69 (0.58-0.81) | 0.0981 | 1 |
|  | N.A. | 561 (0.4%) | 47 (0.05%) | 0.2 (0.17-0.24) | < 0.001 | < 0.001 | 0 (0-6.47E+115) | 1 | 1 |
|  | Unknown | 89345 (64.28%) | 65220 (67.54%) | 1.76 (1.73-1.79) | < 0.001 | < 0.001 | 0.73 (0.66-0.8) | < 0.05 | 0.3423 |
| Fever |  |  |  |  |  |  |  |  |  |
|  | No | 27543 (19.82%) | 20773 (21.51%) | Reference | Reference | Reference | Reference | Reference | Reference |
|  | Yes | 98796 (71.08%) | 60051 (62.19%) | 0.81 (0.79-0.82) | < 0.001 | < 0.001 | 0.95 (0.85-1.05) | 0.4048 | 1 |
|  | Unknown | 12649 (9.1%) | 15743 (16.3%) | 1.65 (1.61-1.69) | < 0.001 | < 0.001 | 1.28 (1.02-1.6) | 0.1419 | 1 |
| Cough |  |  |  |  |  |  |  |  |  |
|  | No | 21042 (15.14%) | 17058 (17.66%) | Reference | Reference | Reference | Reference | Reference | Reference |
|  | Yes | 106745 (76.8%) | 64660 (66.96%) | 0.75 (0.73-0.76) | < 0.001 | < 0.001 | 0.81 (0.72-0.91) | < 0.05 | 0.4745 |
|  | Unknown | 11201 (8.06%) | 14849 (15.38%) | 1.64 (1.59-1.68) | < 0.001 | < 0.001 | 1.02 (0.8-1.3) | 0.8928 | 1 |
| Sore throat |  |  |  |  |  |  |  |  |  |
|  | No | 74822 (53.83%) | 49699 (51.47%) | Reference | Reference | Reference | Reference | Reference | Reference |
|  | Yes | 31003 (22.31%) | 15114 (15.65%) | 0.73 (0.72-0.75) | < 0.001 | < 0.001 | 1.1 (0.97-1.25) | 0.4388 | 1 |
|  | Unknown | 33163 (23.86%) | 31754 (32.88%) | 1.44 (1.42-1.47) | < 0.001 | < 0.001 | 1.01 (0.79-1.28) | 0.9714 | 1 |
| Dyspnea |  |  |  |  |  |  |  |  |  |
|  | No | 32928 (23.69%) | 11309 (11.71%) | Reference | Reference | Reference | Reference | Reference | Reference |
|  | Yes | 89491 (64.39%) | 73149 (75.75%) | 2.38 (2.33-2.43) | < 0.001 | < 0.001 | 1.42 (1.26-1.62) | < 0.001 | < 0.001 |
|  | Unknown | 16569 (11.92%) | 12109 (12.54%) | 2.13 (2.07-2.19) | < 0.001 | < 0.001 | 1.24 (0.97-1.59) | 0.1569 | 1 |
| Respiratory distress |  |  |  |  |  |  |  |  |  |
|  | No | 43642 (31.4%) | 17237 (17.85%) | Reference | Reference | Reference | Reference | Reference | Reference |
|  | Yes | 69439 (49.96%) | 60529 (62.68%) | 2.21 (2.17-2.25) | < 0.001 | < 0.001 | 1.31 (1.17-1.47) | < 0.001 | < 0.05 |
|  | Unknown | 25907 (18.64%) | 18801 (19.47%) | 1.84 (1.8-1.88) | < 0.001 | < 0.001 | 0.86 (0.69-1.08) | 0.2777 | 1 |
| Oxygen saturation < 95% |  |  |  |  |  |  |  |  |  |
|  | No | 46972 (33.8%) | 15339 (15.88%) | Reference | Reference | Reference | Reference | Reference | Reference |
|  | Yes | 65884 (47.4%) | 63011 (65.25%) | 2.93 (2.88-2.98) | < 0.001 | < 0.001 | 1.32 (1.17-1.48) | < 0.001 | < 0.05 |
|  | Unknown | 26132 (18.8%) | 18217 (18.86%) | 2.14 (2.09-2.18) | < 0.001 | < 0.001 | 1.08 (0.86-1.34) | 0.5858 | 1 |
| Diarrhea |  |  |  |  |  |  |  |  |  |
|  | No | 81912 (58.93%) | 53323 (55.22%) | Reference | Reference | Reference | Reference | Reference | Reference |
|  | Yes | 20931 (15.06%) | 10122 (10.48%) | 0.74 (0.73-0.76) | < 0.001 | < 0.001 | 1.02 (0.88-1.17) | 1 | 1 |
|  | Unknown | 36145 (26.01%) | 33122 (34.3%) | 1.41 (1.39-1.43) | < 0.001 | < 0.001 | 0.94 (0.69-1.29) | 1 | 1 |
| Vomiting |  |  |  |  |  |  |  |  |  |
|  | No | 88567 (63.72%) | 55916 (57.9%) | Reference | Reference | Reference | Reference | Reference | Reference |
|  | Yes | 11477 (8.26%) | 6275 (6.5%) | 0.87 (0.84-0.89) | < 0.001 | < 0.001 | 0.99 (0.84-1.18) | 1 | 1 |
|  | Unknown | 38944 (28.02%) | 34376 (35.6%) | 1.4 (1.38-1.42) | < 0.001 | < 0.001 | 0.89 (0.65-1.22) | 1 | 1 |
| Risk Factor |  |  |  |  |  |  |  |  |  |
|  | No | 63309 (45.55%) | 26390 (27.33%) | Reference | Reference | Reference | Reference | Reference | Reference |
|  | Yes | 75679 (54.45%) | 70177 (72.67%) | 2.23 (2.19-2.26) | < 0.001 | < 0.001 | 1.08 (0.89-1.31) | 0.4298 | 1 |
| Postpartum period† |  |  |  |  |  |  |  |  |  |
|  | No | 126458 (90.98%) | 85701 (88.75%) | Reference | Reference | Reference | Reference | Reference | Reference |
|  | Yes | 484 (0.35%) | 137 (0.14%) | 0.42 (0.36-0.49) | < 0.001 | < 0.001 | 0.54 (0.21-1.39) | 0.3832 | 1 |
|  | Unknown | 12046 (8.67%) | 10729 (11.11%) | 1.31 (1.28-1.35) | < 0.001 | < 0.001 | 1.19 (0.96-1.48) | 0.3832 | 1 |
| Chronic Cardiovascular Disease |  |  |  |  |  |  |  |  |  |
|  | No | 86280 (62.08%) | 43873 (45.43%) | Reference | Reference | Reference | Reference | Reference | Reference |
|  | Yes | 37500 (26.98%) | 38416 (39.78%) | 2.02 (1.98-2.05) | < 0.001 | < 0.001 | 0.96 (0.85-1.09) | 0.6098 | 1 |
|  | Unknown | 15208 (10.94%) | 14278 (14.79%) | 1.85 (1.81-1.89) | < 0.001 | < 0.001 | 1.28 (1.02-1.62) | 0.1586 | 1 |
| Chronic Hematologic Disease |  |  |  |  |  |  |  |  |  |
|  | No | 107620 (77.43%) | 65069 (67.38%) | Reference | Reference | Reference | Reference | Reference | Reference |
|  | Yes | 911 (0.66%) | 992 (1.03%) | 1.8 (1.67-1.94) | < 0.001 | < 0.001 | 1.81 (1.08-3.04) | 0.1175 | 1 |
|  | Unknown | 30457 (21.91%) | 30506 (31.59%) | 1.66 (1.63-1.68) | < 0.001 | < 0.001 | 0.82 (0.43-1.58) | 0.6175 | 1 |
| Down Syndrome |  |  |  |  |  |  |  |  |  |
|  | No | 108151 (77.81%) | 65700 (68.04%) | Reference | Reference | Reference | Reference | Reference | Reference |
|  | Yes | 271 (0.19%) | 300 (0.31%) | 1.82 (1.59-2.09) | < 0.001 | < 0.001 | 0.84 (0.37-1.89) | 1 | 1 |
|  | Unknown | 30566 (21.99%) | 30567 (31.65%) | 1.65 (1.62-1.67) | < 0.001 | < 0.001 | 1.28 (0.64-2.56) | 1 | 1 |
| Chronic Hepatic Disease |  |  |  |  |  |  |  |  |  |
|  | No | 107413 (77.28%) | 64700 (67%) | Reference | Reference | Reference | Reference | Reference | Reference |
|  | Yes | 841 (0.61%) | 1246 (1.29%) | 2.46 (2.29-2.65) | < 0.001 | < 0.001 | 1.8 (1.17-2.77) | < 0.05 | 1 |
|  | Unknown | 30734 (22.11%) | 30621 (31.71%) | 1.65 (1.63-1.68) | < 0.001 | < 0.001 | 1.68 (0.96-2.92) | 0.1282 | 1 |
| Asthma |  |  |  |  |  |  |  |  |  |
|  | No | 105508 (75.91%) | 64320 (66.61%) | Reference | Reference | Reference | Reference | Reference | Reference |
|  | Yes | 3980 (2.86%) | 1984 (2.05%) | 0.82 (0.78-0.86) | < 0.001 | < 0.001 | 0.82 (0.61-1.09) | 0.5067 | 1 |
|  | Unknown | 29500 (21.22%) | 30263 (31.34%) | 1.68 (1.66-1.71) | < 0.001 | < 0.001 | 0.96 (0.62-1.49) | 0.8847 | 1 |
| Diabetes Mellitus |  |  |  |  |  |  |  |  |  |
|  | No | 91270 (65.67%) | 48396 (50.12%) | Reference | Reference | Reference | Reference | Reference | Reference |
|  | Yes | 28364 (20.41%) | 30400 (31.48%) | 2.02 (1.99-2.06) | < 0.001 | < 0.001 | 1.19 (1.06-1.35) | < 0.05 | 1 |
|  | Unknown | 19354 (13.92%) | 17771 (18.4%) | 1.73 (1.7-1.77) | < 0.001 | < 0.001 | 0.76 (0.61-0.95) | < 0.05 | 1 |
| Chronic Neurological Disease |  |  |  |  |  |  |  |  |  |
|  | No | 106170 (76.39%) | 62073 (64.28%) | Reference | Reference | Reference | Reference | Reference | Reference |
|  | Yes | 3326 (2.39%) | 5745 (5.95%) | 2.95 (2.85-3.07) | < 0.001 | < 0.001 | 1.99 (1.58-2.5) | < 0.001 | < 0.001 |
|  | Unknown | 29492 (21.22%) | 28749 (29.77%) | 1.67 (1.64-1.69) | < 0.001 | < 0.001 | 1.72 (1.22-2.44) | < 0.05 | 1 |
| Chronic Pneumopathy |  |  |  |  |  |  |  |  |  |
|  | No | 106011 (76.27%) | 62294 (64.51%) | Reference | Reference | Reference | Reference | Reference | Reference |
|  | Yes | 3314 (2.38%) | 5220 (5.41%) | 2.68 (2.58-2.78) | < 0.001 | < 0.001 | 1.38 (1.1-1.73) | < 0.05 | 1 |
|  | Unknown | 29663 (21.34%) | 29053 (30.09%) | 1.67 (1.64-1.69) | < 0.001 | < 0.001 | 0.64 (0.43-0.96) | 0.0684 | 1 |
| Immunodeficiency / Immunosuppression |  |  |  |  |  |  |  |  |  |
|  | No | 105999 (76.26%) | 63113 (65.36%) | Reference | Reference | Reference | Reference | Reference | Reference |
|  | Yes | 2965 (2.13%) | 3495 (3.62%) | 1.98 (1.9-2.06) | < 0.001 | < 0.001 | 2.48 (1.92-3.21) | < 0.001 | < 0.001 |
|  | Unknown | 30024 (21.6%) | 29959 (31.02%) | 1.68 (1.65-1.7) | < 0.001 | < 0.001 | 0.82 (0.54-1.25) | 0.4432 | 1 |
| Chronic Renal Disease |  |  |  |  |  |  |  |  |  |
|  | No | 105644 (76.01%) | 61497 (63.68%) | Reference | Reference | Reference | Reference | Reference | Reference |
|  | Yes | 3522 (2.53%) | 6298 (6.52%) | 3.07 (2.96-3.18) | < 0.001 | < 0.001 | 1.49 (1.19-1.86) | < 0.05 | 0.7342 |
|  | Unknown | 29822 (21.46%) | 28772 (29.79%) | 1.66 (1.63-1.68) | < 0.001 | < 0.001 | 0.75 (0.52-1.08) | 0.1981 | 1 |
| Obesity |  |  |  |  |  |  |  |  |  |
|  | No | 102374 (73.66%) | 60824 (62.99%) | Reference | Reference | Reference | Reference | Reference | Reference |
|  | Yes | 7703 (5.54%) | 6237 (6.46%) | 1.36 (1.32-1.4) | < 0.001 | < 0.001 | 1.16 (0.96-1.39) | 0.3912 | 1 |
|  | Unknown | 28911 (20.8%) | 29506 (30.55%) | 1.72 (1.69-1.75) | < 0.001 | < 0.001 | 1.13 (0.87-1.48) | 0.4385 | 1 |
| High Blood Pressure |  |  |  |  |  |  |  |  |  |
|  | No | 63309 (45.55%) | 26390 (27.33%) | Reference | Reference | Reference | Reference | Reference | Reference |
|  | Yes | 14661 (10.55%) | 14356 (14.87%) | 2.35 (2.3-2.4) | < 0.001 | < 0.001 | 1.01 (0.85-1.2) | 0.8998 | 1 |
|  | Unknown | 61018 (43.9%) | 55821 (57.81%) | 2.2 (2.16-2.23) | < 0.001 | < 0.001 | 1.08 (0.92-1.27) | 0.8596 | 1 |
| Unlisted risk factors |  |  |  |  |  |  |  |  |  |
|  | No | 99739 (71.76%) | 57177 (59.21%) | Reference | Reference | Reference | Reference | Reference | Reference |
|  | Yes | 18154 (13.06%) | 19216 (19.9%) | 1.85 (1.81-1.88) | < 0.001 | < 0.001 | 1.12 (0.98-1.26) | 0.3025 | 1 |
|  | Unknown | 21095 (15.18%) | 20174 (20.89%) | 1.67 (1.64-1.7) | < 0.001 | < 0.001 | 1.07 (0.89-1.29) | 0.5569 | 1 |
| Ventilatory Support |  |  |  |  |  |  |  |  |  |
|  | No | 47265 (34.01%) | 10485 (10.86%) | Reference | Reference | Reference | Reference | Reference | Reference |
|  | Yes, invasive | 7248 (5.21%) | 36399 (37.69%) | 22.64 (22.1-23.19) | < 0.001 | < 0.001 | 16.23 (14.05-18.75) | < 0.001 | < 0.001 |
|  | Yes, non-invasive | 60862 (43.79%) | 27557 (28.54%) | 2.04 (2-2.08) | < 0.001 | < 0.001 | 1.45 (1.3-1.62) | < 0.001 | < 0.001 |
|  | Unknown | 23613 (16.99%) | 22126 (22.91%) | 4.22 (4.14-4.31) | < 0.001 | < 0.001 | 2.74 (2.27-3.31) | < 0.001 | < 0.001 |
| X-ray |  |  |  |  |  |  |  |  |  |
|  | Normal | 3692 (2.66%) | 1483 (1.54%) | Reference | Reference | Reference | Reference | Reference | Reference |
|  | Interstitial infiltrate | 23453 (16.87%) | 16027 (16.6%) | 1.7 (1.65-1.76) | < 0.001 | < 0.001 | 1.31 (1.05-1.64) | 0.4185 | 1 |
|  | Consolidation | 3173 (2.28%) | 2543 (2.63%) | 2 (1.91-2.08) | < 0.001 | < 0.001 | 1.36 (1.03-1.78) | 0.4185 | 1 |
|  | Mixed | 3815 (2.74%) | 3089 (3.2%) | 2.02 (1.94-2.1) | < 0.001 | < 0.001 | 1.92 (1.48-2.49) | 0.0567 | 1 |
|  | Other | 24473 (17.61%) | 13255 (13.73%) | 1.35 (1.3-1.4) | < 0.001 | < 0.001 | 1.72 (1.37-2.16) | 0.0685 | 1 |
|  | Unrealized | 25804 (18.57%) | 14880 (15.41%) | 1.44 (1.39-1.49) | < 0.001 | < 0.001 | 1.49 (1.19-1.85) | 0.2419 | 1 |
|  | Unknown | 54578 (39.27%) | 45290 (46.9%) | 2.07 (2-2.13) | < 0.001 | < 0.001 | 1.39 (1.12-1.74) | 0.3585 | 1 |
| Computerized tomography |  |  |  |  |  |  |  |  |  |
|  | No | 48 (0.66%) | 14 (0.25%) | Reference | Reference | Reference | Reference | Reference | Reference |
|  | COVID Typical | 3672 (50.14%) | 2603 (46.87%) | 2.43 (1.77-3.33) | < 0.05 | 0.7437 | 2.22 (1.36-3.63) | 0.1868 | 1 |
|  | COVID Indetermined | 136 (1.86%) | 147 (2.65%) | 3.71 (2.64-5.2) | < 0.001 | < 0.05 | 2.83 (1.68-4.76) | 0.1148 | 1 |
|  | COVID Atypical | 63 (0.86%) | 73 (1.31%) | 3.97 (2.77-5.7) | < 0.001 | < 0.05 | 3.93 (2.27-6.82) | 0.06 | 1 |
|  | Other | 399 (5.45%) | 252 (4.54%) | 2.17 (1.56-3) | < 0.05 | 1 | 1.9 (1.15-3.16) | 0.1883 | 1 |
|  | Unrealized | 2010 (27.45%) | 1567 (28.21%) | 2.67 (1.95-3.67) | < 0.05 | 0.404 | 3.03 (1.85-4.96) | 0.0807 | 1 |
|  | Unknown | 995 (13.59%) | 898 (16.17%) | 3.09 (2.25-4.26) | < 0.001 | 0.0982 | 3.35 (2.04-5.5) | 0.06 | 1 |
| Intensive Care Unit |  |  |  |  |  |  |  |  |  |
|  | No | 87797 (63.17%) | 30492 (31.58%) | Reference | Reference | Reference | Reference | Reference | Reference |
|  | Yes | 26166 (18.83%) | 45024 (46.62%) | 4.96 (4.87-5.04) | < 0.001 | < 0.001 | 3.14 (2.82-3.48) | < 0.001 | < 0.001 |
|  | Unknown | 25025 (18.01%) | 21051 (21.8%) | 2.42 (2.38-2.47) | < 0.001 | < 0.001 | 1.87 (1.48-2.37) | < 0.001 | < 0.001 |
| Previous grippal case** |  |  |  |  |  |  |  |  |  |
|  | No | 71856 (51.7%) | 41419 (42.89%) | Reference | Reference | Reference | Reference | Reference | Reference |
|  | Yes | 29837 (21.47%) | 24204 (25.06%) | 1.41 (1.38-1.43) | < 0.001 | < 0.001 | 1.32 (1.19-1.47) | < 0.001 | < 0.05 |
|  | Unknown | 37295 (26.83%) | 30944 (32.04%) | 1.44 (1.42-1.46) | < 0.001 | < 0.001 | 1.39 (1.22-1.59) | < 0.001 | < 0.05 |
| Age Range |  |  |  |  |  |  |  |  |  |
|  | (20,30] | 9784 (7.04%) | 1158 (1.2%) | Reference | Reference | Reference | Reference | Reference | Reference |
|  | (0,1] | 329 (0.24%) | 30 (0.03%) | 0.77 (0.68-0.88) | 0.3539 | 1 | 0 (0-3.846E+76) | 0.9646 | 1 |
|  | (1,6] | 853 (0.61%) | 74 (0.08%) | 0.73 (0.67-0.8) | < 0.05 | 1 | 0.19 (0.11-0.35) | 0.2403 | 1 |
|  | (6,20] | 2669 (1.92%) | 342 (0.35%) | 1.08 (1.04-1.13) | 0.3539 | 1 | 0.44 (0.32-0.59) | 0.2403 | 1 |
|  | (30,40] | 21726 (15.63%) | 3603 (3.73%) | 1.4 (1.37-1.44) | < 0.001 | < 0.001 | 0.85 (0.74-0.99) | 0.9151 | 1 |
|  | (40,50] | 26739 (19.24%) | 7328 (7.59%) | 2.32 (2.26-2.37) | < 0.001 | < 0.001 | 1.69 (1.48-1.94) | < 0.05 | 1 |
|  | (50,60] | 28348 (20.4%) | 13753 (14.24%) | 4.1 (4.01-4.19) | < 0.001 | < 0.001 | 2.09 (1.83-2.38) | < 0.001 | 0.103 |
|  | (60,70] | 23951 (17.23%) | 22724 (23.53%) | 8.02 (7.84-8.19) | < 0.001 | < 0.001 | 4.22 (3.7-4.81) | < 0.001 | < 0.001 |
|  | (70,80] | 15653 (11.26%) | 24477 (25.35%) | 13.21 (12.92-13.51) | < 0.001 | < 0.001 | 7.37 (6.45-8.41) | < 0.001 | < 0.001 |
|  | (80,90] | 7547 (5.43%) | 18099 (18.74%) | 20.26 (19.8-20.73) | < 0.001 | < 0.001 | 12.68 (11.07-14.53) | < 0.001 | < 0.001 |
|  | > 90 | 1389 (1%) | 4979 (5.16%) | 30.29 (29.41-31.19) | < 0.001 | < 0.001 | 28.91 (24.5-34.11) | < 0.001 | < 0.001 |
| Illness onset to hospitalization (days) |  |  |  |  |  |  |  |  |  |
|  | (0,3] | 23532 (18.66%) | 20681 (24.4%) | Reference | Reference | Reference | Reference | Reference | Reference |
|  | (3,6] | 30246 (23.99%) | 19545 (23.06%) | 0.74 (0.72-0.75) | < 0.001 | < 0.001 | 0.94 (0.85-1.05) | 0.484 | 1 |
|  | (6,9] | 31096 (24.66%) | 15396 (18.16%) | 0.56 (0.55-0.57) | < 0.001 | < 0.001 | 0.67 (0.59-0.75) | < 0.001 | < 0.05 |
|  | > 9 | 30502 (24.19%) | 14862 (17.53%) | 0.55 (0.55-0.56) | < 0.001 | < 0.001 | 0.59 (0.51-0.68) | < 0.001 | < 0.05 |
|  | Unknown*** | 10717 (8.5%) | 14290 (16.86%) | 1.52 (1.49-1.55) | < 0.001 | < 0.001 | 1.89 (1.66-2.15) | < 0.001 | < 0.001 |
| Illness onset to cure or death (days) |  |  |  |  |  |  |  |  |  |
|  | (0,9] | 26703 (21.23%) | 30781 (32.02%) | Reference | Reference | Reference | Reference | Reference | Reference |
|  | (9,14] | 35358 (28.11%) | 20226 (21.04%) | 0.5 (0.49-0.5) | < 0.001 | < 0.001 | 0.79 (0.7-0.89) | < 0.05 | 1 |
|  | (14,20] | 34176 (27.17%) | 18046 (18.77%) | 0.46 (0.45-0.47) | < 0.001 | < 0.001 | 0.82 (0.7-0.96) | 0.3089 | 1 |
|  | > 20 | 29306 (23.3%) | 24077 (25.04%) | 0.71 (0.7-0.72) | < 0.001 | < 0.001 | 0.96 (0.78-1.19) | 1 | 1 |
|  | Unknown*** | 255 (0.2%) | 3008 (3.13%) | 10.23 (9.41-11.13) | < 0.001 | < 0.001 | 0.79 (0.39-1.56) | 1 | 1 |
| Hospitalization duration (days) |  |  |  |  |  |  |  |  |  |
|  | (0,4] | 31523 (26.99%) | 21656 (25.64%) | Reference | Reference | Reference | Reference | Reference | Reference |
|  | (4,7] | 29323 (25.11%) | 13773 (16.31%) | 0.68 (0.67-0.7) | < 0.001 | < 0.001 | 0.67 (0.6-0.74) | < 0.001 | < 0.001 |
|  | (7,13] | 30220 (25.87%) | 19743 (23.38%) | 0.95 (0.94-0.97) | < 0.001 | < 0.05 | 0.86 (0.76-0.98) | 0.2506 | 1 |
|  | > 13 | 23819 (20.39%) | 23125 (27.38%) | 1.41 (1.39-1.44) | < 0.001 | < 0.001 | 0.84 (0.7-1) | 0.2506 | 1 |
|  | Unknown*** | 1908 (1.63%) | 6162 (7.3%) | 4.7 (4.54-4.87) | < 0.001 | < 0.001 | 4.56 (3.69-5.64) | < 0.001 | < 0.001 |

Death and cure proportions represented by n (%). Schooling level expressed as the number of years of study. Adj=Adjusted. \*P-values were corrected by the number of comparisons with the reference level by the Holm-Sidak method. †Odds Ratios were adjusted by all significant variables in bivariate models (OR) and added as confounding variables in the multivariate logistic models (aOR) whenever applicable. ‡Up to 45 days after giving birth. \*\*Diagnosed and notified as COVID-19 before hospitalization. \*\*\*Zero included in the unknown.

**Figure 1:**
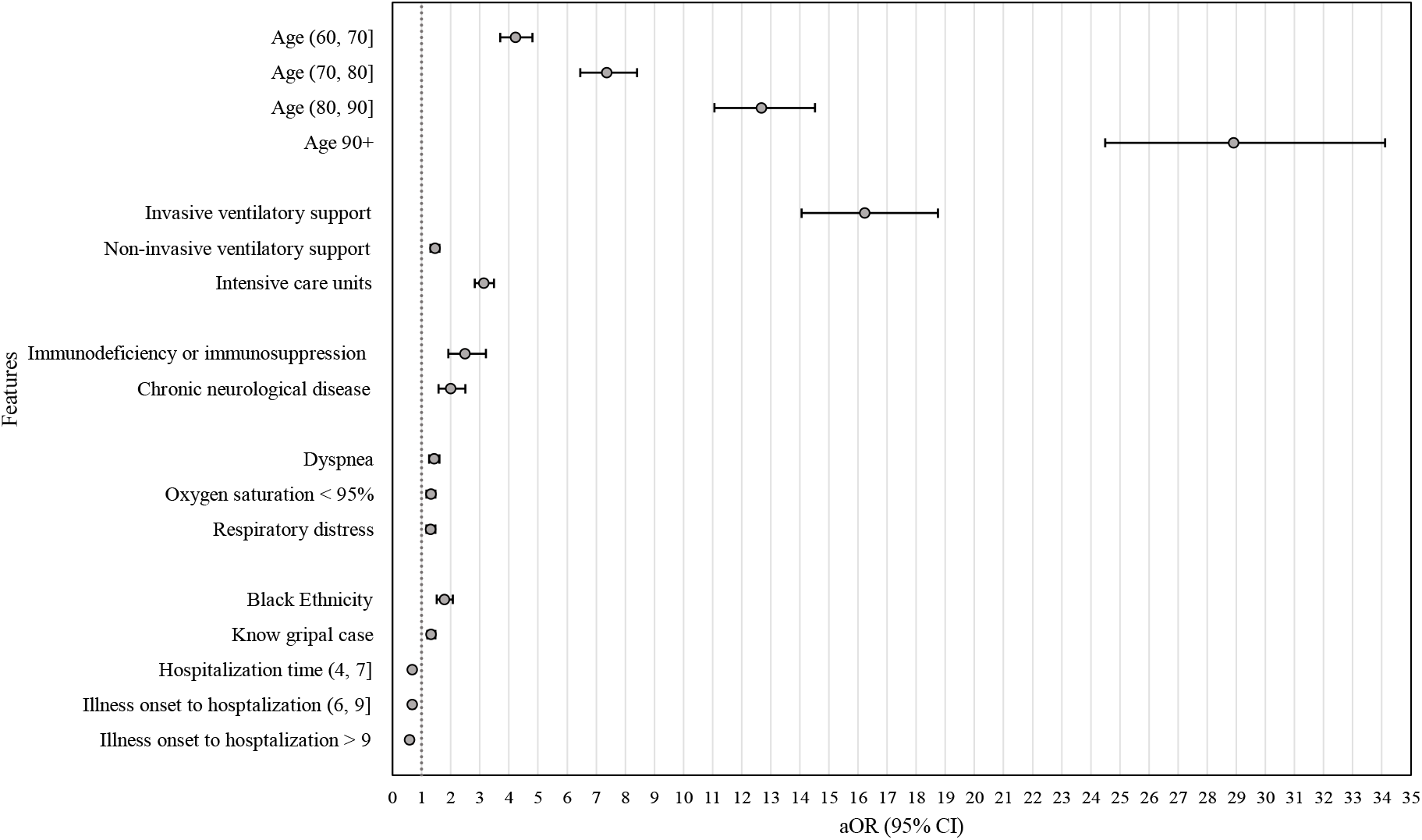
Risk factors for COVID-19 death. For each feature, aOR was estimated against its reference (Table 3), its mean represented by the dot and the 95% CI given by the respective black line. Vertical dotted line at aOR = 1. All significant variables in bivariate models were added as confounding variables in the multivariate logistic models.

The combination of oxygen saturation < 95%, dyspnea, and respiratory distress was shown to be the worst association of respiratory symptoms with death, followed by oxygen saturation < 95% with respiratory distress, oxygen saturation < 95% with dyspnea, and dyspnea with respiratory distress (Figure 2; appendix p 1).

**Figure 2:**
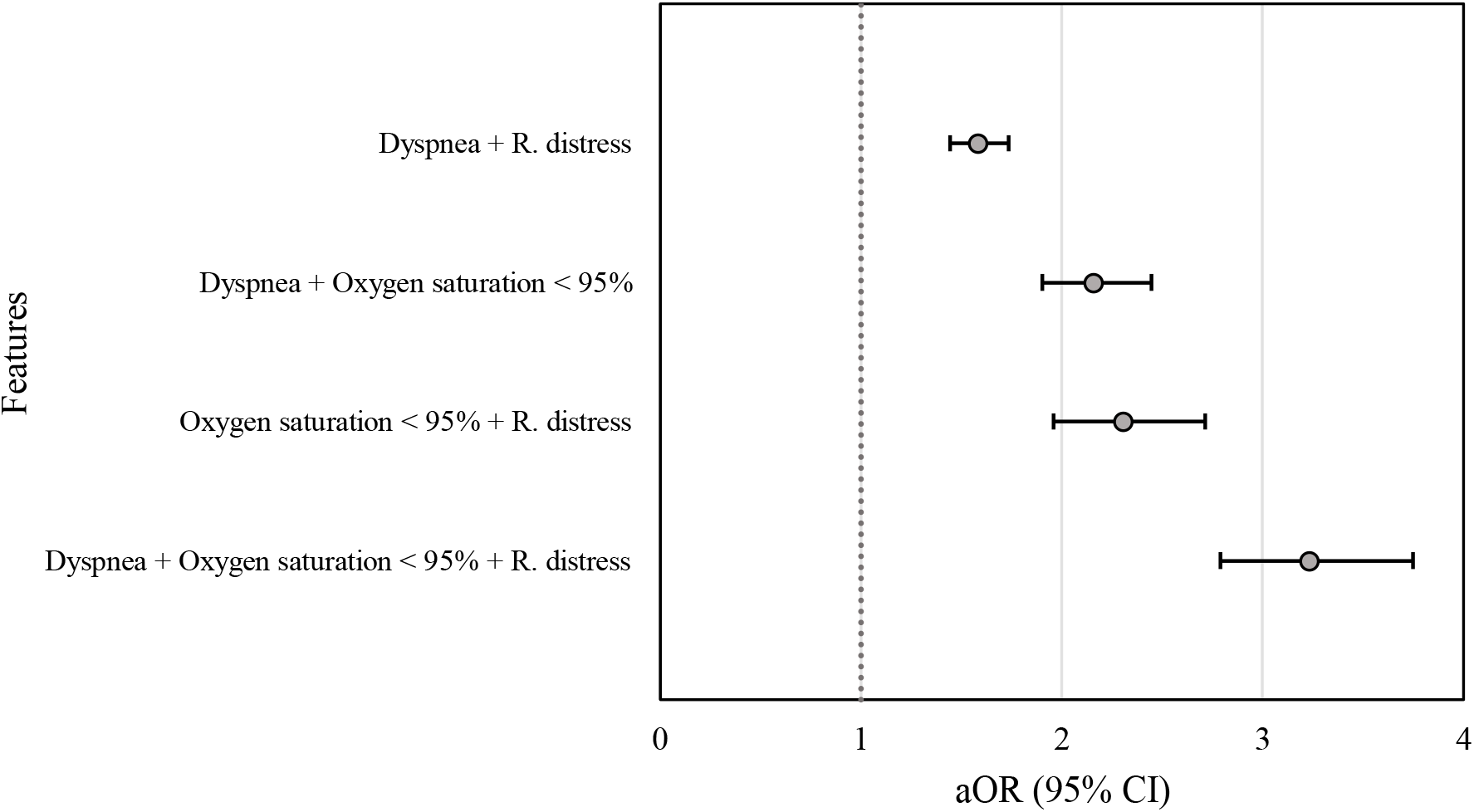
Risk of the combination of respiratory symptoms for COVID-19 death. For each respiratory symptoms combination, the aOR was estimated against subjects without all three respiratory symptoms (appendix p 1), its mean represented by the dot and the 95% CI given by the respective black line. Vertical dotted line at aOR = 1. The confounding variables, i.e., ethnicity, ventilatory support, and intensive care unit, were kept in equal proportions in the multivariate logistic models. R. distress=Respiratory distress.

Regarding the impact of having one risk factor alone, patients aged (0, 20] had the worst outcome, followed by those (20, 40], and those (40, 60] (Figure 3; appendix p 1). To what concerns the impact of any two or more risk factors, patients aged (20, 40] had the worst outcome, followed by those (40, 60], and minor impact in (60, 80] (Figure 3; appendix p 2).

**Figure 3:**
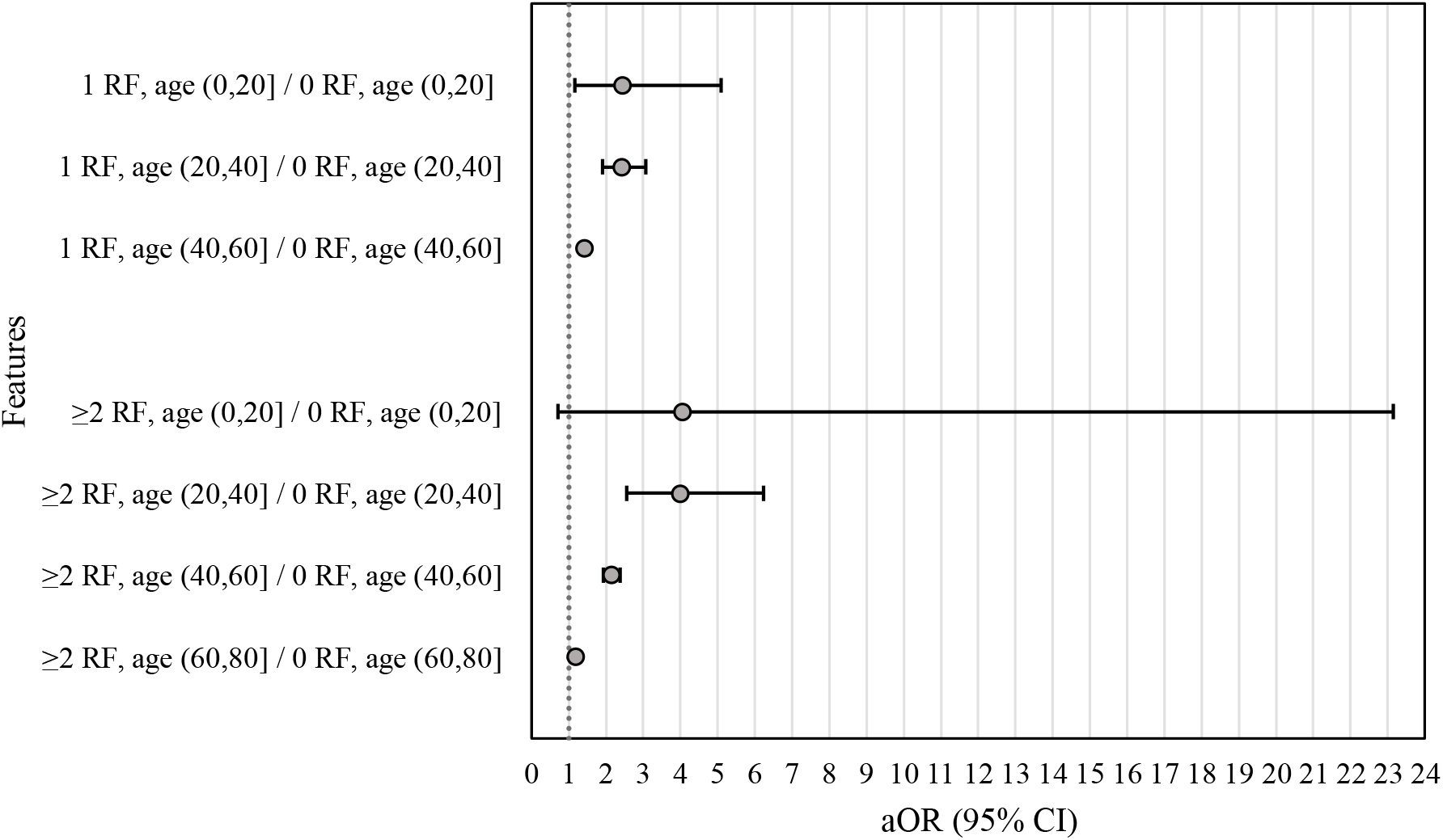
Risk of the combination of the number of any COVID-19 risk factors with age for death. For each number of risk factors and age combination, the aOR was estimated against its reference (appendix p 2), its mean represented by the dot and the 95% CI given by the respective black line. Vertical dotted line at aOR = 1. The confounding variables, i.e., ethnicity, ventilatory support, and intensive care unit, were kept in equal proportions in the multivariate logistic models. RF= Risk Factor.

Finally, we analyzed the impact of diabetes mellitus (DM) combined with other risk factors on death, focusing on different age intervals. In patients up to 20 years old, DM, either with chronic hematologic diseases or obesity, leads to approximately a twenty-one-fold increase in death risk, followed by roughly a six-fold by brown ethnicity (white as reference) (Figure 4; appendix p 3). At age (20, 40], DM, either with chronic cardiovascular diseases or chronic high blood pressure, showed a four-fold increase in death risk (Figure 4; appendix p 3). Yet, among the younger population, DM, when associated either with chronic neurological diseases, chronic pneumopathy diseases, chronic renal diseases, or unlisted risk factors, shows a recurring pattern of worse outcome (Figure 4).

**Figure 4:**
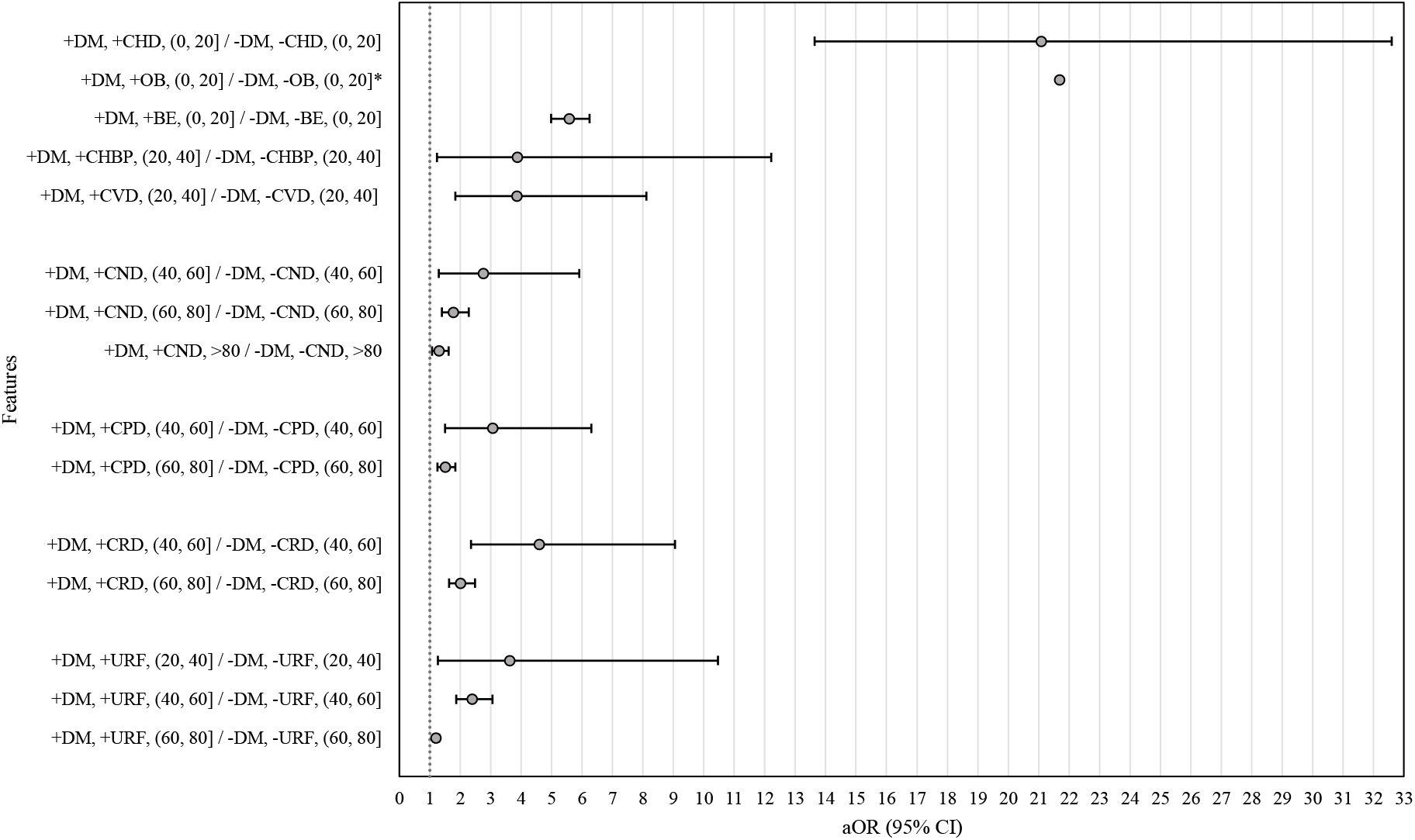
Risk of the combination of diabetes mellitus and other risk factors for COVID-19 death. For each number of risk factors and age combination, the aOR was estimated against its reference (appendix p 3), its mean represented by the dot and the 95% CI given by the respective black line. Vertical dotted line at aOR = 1. The confounding variables, i.e., ethnicity, ventilatory support, and intensive care unit, were kept in equal proportions in the multivariate logistic models. * 95% CI not calculated. DM=Diabetes Mellitus. CHD=Chronic Hematologic Disease. OB=Obesity. BE=Brown Ethnicity. CHBP=Chronic High Blood Pressure. CVD=Chronic Vascular Disease. CND=Chronic Neurologic Disease. CPD=Chronic Pneumopathy Disease. CRD=Chronic Renal Disease. UR =‘Unlisted Risk actors’.

## DISCUSSION

This large-scale retrospective cohort study identified and estimated the impact of several risk factors for death in COVID-19 Brazilian hospitalized patients, as well as their severe increase in association with diabetes. Individually, advanced age, invasive ventilatory support, admission to intensive care units, and multiple respiratory symptoms were associated with higher death odds. Usage of non-invasive ventilatory support, immunodeficiency or immunosuppression, black ethnicity, and COVID-19 diagnosis previous to hospitalization were also aggravating factors for death, with less impact. Younger diabetic patients either self-declared as brown ethnicity or with some comorbidities, mainly chronic hematologic disease and obesity, presented an outstanding death risk. As protective factors, we found hospitalization duration of (4, 7] and illness onset to hospitalization interval greater than 6 days.

Fever and cough have been reported as the most prevalent COVID-19 symptoms worldwide, with substantial heterogeneity between countries.^7,18,19^ Our study of hospitalized patients confirmed this fact, with fever reported in 67·4% of cases and cough in 72·8%. Compared with other countries, the prevalence found in our data was among the highest for both symptoms, with 32% in Korea and 83% in Singapore for fever and 18% in Korea and 76% in the Netherlands for cough.^18^ Although high prevalence, no statistically significant impact on death was found here in Brazil. Many other countries, such as Italy,^20^ Spain,^21^ and Egypt,^22^ found a correlation of dyspnea or low oxygen saturation with higher hospital deaths. Our data corroborate this, showing that a combination of both symptoms more than doubles hospital death risk. Additionally, respiratory distress and the association of multiple respiratory symptoms are important risk factors, with Brazilian patients manifesting all three respiratory symptoms having an approximately three-fold increase in death risk.

Regarding the usage of intensive care units, invasive ventilatory support and non-invasive ventilatory support, our data shows respectively 30·2%, 18·5% and 37·5%, higher than the respectively formerly reported 17%, 9% and 19% among worldwide hospitalized patients.^18^ Unfortunately, almost half of COVID-19 Brazilian hospitalized patients are only diagnosed during admission. Despite one of the countries with most cases, Brazil is in 125^th^ place in the number of SARS-CoV-2 real-time polymerase chain reaction exams per million inhabitants.^23^ We suggest that the high usage of intensive care units and ventilatory support in Brazil, compared to elsewhere, may be in part due to late COVID-19 diagnosis. This fact reflects significant disparities in health care accessibility and socioeconomic inequalities throughout the country.^24^ Hospitalization duration of (4, 7] and illness onset to hospitalization interval greater than 6 days as protective factors are probably related to viral shedding. Better understanding of this dynamic is a must for optimizing pandemic hospital management in Brazil and saving lives, especially for risk groups composed of subpopulations.

Aging leads to a natural decline in the immune system and a chronic inflammatory state,^25^ and has been reported as a biological vulnerability for COVID-19 mortality.^21,26,27^ We also found age greater than 60 as a prominent risk factor, particularly over 90 years. A nationwide large cohort study in the United Kingdom showed that while age is per se an independent risk factor, the mortality on elderly patients is partially explained by the higher prevalence of comorbidities, such as poorer lung function, hypertension, muscle weakness, and multiple long-term conditions.^27^ Conversely, our data revealed that the impact of having one or a combination of two or more risk factors on mortality are progressively higher in ages (60, 80], (40, 60], (20, 40], and (0, 20] compared with people of the same age interval without comorbidities. This discrepancy could be due to the British study using, as a reference group, people less than 65 years old with or without comorbidities. The synergism of COVID-19, pro-inflammatory comorbidities and a young immune profile, leading to overproduction of cytokines, must be further investigated as it partially explains the higher mortality in this intersection group.

Beyond advanced age, some features, as ethnicity and gender, have been also identified as biological concern for COVID-19 mortality.^28,29^ Although over half of the hospitalized patients were men, gender independently was not statistically associated with higher hospital death risk in Brazil, despite other reports which associate male sex with worse in-hospital outcomes.^30^ Still, biological factors underlying higher mortality in ethnic groups are limited and need to be further addressed. We observed young brown individuals with diabetes and black ethnicity as population subgroups at higher risk for severe COVID-19. A longitudinal analysis of 2012–16 economic recession in Brazil showed an increase in mortality in the same groups, due to socioeconomic factors, labour market participation, and underlying morbidities.^31^ As in past economic crises, actual pandemics highlights Brazilian social and health care access inequalities. Historically, compared with white, black and brown Brazilians have lower schooling levels, a higher index of informal jobs, lower incomes, and restricted access to the health care system,^32^ particularly for men.^33^ Other study showed that COVID-19 death rate is higher in these Brazilians subpopulations and might be partly due to non-ICU admission, especially for brown.^34^ We also found brown ethnicity as a independent risk factor prior to using ventilatory support and ICU usage as confounding variables, what supports the former study hypothesis.^34^

Diabetes Mellitus is often associated with other comorbidities and has been consistently correlated with an increase in severity and mortality of COVID-19 patients.^14,35^ Diabetes, high blood pressure, chronic renal diseases, Alzheimer disease, pneumopathy, and obesity are conditions that lead to an upregulation expression of ACE2, resulting in increased viral uptake, turning them into susceptible sites.^36,37^ ACE2 (Angiotensin-converting enzyme 2) is one of the surface receptors used by SARS-CoV-2 to enter host cells, and it is constitutively expressed in a wide variety of epithelial, non-epithelial cells, and organs as lungs, heart, kidneys, and adipocyte tissue.^36,38^ Those tissues normally express higher amounts of ACE2 receptor, added to the fact that some conditions potentially increased ACE2 expression. This combination partially explains higher death risk in COVID-19 patients with comorbidities. The infection itself can cause deleterious effects on immunity, and a synergic event between diseases causes hyperactivity of the immune system, leading to a chronic proinflammatory state, what contributes to the cytokine storm seen in some severe COVID-19 patients.^14,36^

Our study has some key limitations, mainly regarding data quality, annotation accuracy, suspected under-reported data, and lack of government open-access information of non-hospitalized patients in Brazil. This is a relevant bias as studies based only on hospitalized patients are focused on more severe cases and fatalities. Furthermore, we cannot guarantee that missing information is not subject to bias. Although extensive, some fundamental information that greatly affects the prognosis of the disease is missing in the SRAG form, mainly regarding smoking, alcohol intake, oncological disorders, and chronic high blood pressure.^39–42^ As for our study’s relevance, drawing attention to diabetes, lack of reliable data on Body Mass Index (BMI) prevented us from differentiating the impact of obesity and severe obesity in mortality. This is mainly important because obesity is a chronic inflammatory condition associated with cardiometabolic and immune dysfunction, increased risk of diabetes and hematological disease, leading patients to be more susceptible to develop severe forms of COVID-19.^36^ The under-reporting in the BMI may be aggravated due to professionals being not properly instructed on how to use and fill out the form items. Thus, we suggest remodeling the form, together with better orientation on how to deal with information gathering. Nonetheless, despite all Brazilian health system challenges, the average notification delay was 0 days (IQR=2), demonstrating high efficiency in national information flow.

To the best of our knowledge, this is the largest retrospective cohort study of hospitalized Brazilians COVID-19 patients up to date. Older age, multiple respiratory symptoms, younger patients with some risk factor, and younger diabetics with multiple comorbidities are risk groups that play an important role in the death by COVID-19 scenario. Globally, measures have been focused on health workers and older patients as risk groups and priority for vaccination.

Our findings have shone a spotlight upon a risk group that is so far neglected in Brazil, as younger patients with comorbidities and younger diabetic patients with further comorbidities. The understanding of how comorbidities increased risk mortality by COVID-19 is fundamental for implementing appropriate public health and therapeutic strategies. We expect this meaningful nationwide analysis to be highly considered to help establish patient risk stratification upon admission, optimizing hospital management, and guide public policy determinations, including group prioritization for COVID-19 vaccination and vulnerable populations health care accessibility in Brazil.

## Data Availability

DATA SHARING
SRAG data is publicly available on the Ministry of Health of Brazil website. The analysis source code will be available upon request to the authors.

## CONTRIBUTORS

EP conceived the research question. EP, GPA, MRA and MA designed the study and analyses plan. EP obtained and performed initial data cleanup. MRA carried out the analyses. EP and GPA drafted the initial version of the manuscript. All authors drafted and edited the final version of the manuscript. MFFB performed spelling and grammar reviewing. All authors had access to all used data. All authors critically reviewed and approved the final version of the manuscript.

## DATA SHARING

SRAG data is publicly available on the Ministry of Health of Brazil website. The analysis source code will be available upon request to the authors.

## REFERENCES

1 Ludwig S, Zarbock A. Coronaviruses and SARS-CoV-2: A Brief Overview. Anesth Analg 2020; published online April 20. DOI:10.1213/ANE.0000000000004845.

2 Rodriguez-Morales AJ, Gallego V, Escalera-Antezana JP, et al. COVID-19 in Latin America: The implications of the first confirmed case in Brazil. Travel Med Infect Dis 2020; 35: 101613.

3 World Health Organization. WHO Director-General’s statement on IHR Emergency Committee on Novel Coronavirus (2019-nCoV). 2020; published online Jan 30. https://www.who.int/director-general/speeches/detail/who-director-general-s-statement-on-ihr-emergency-committee-on-novel-coronavirus-(2019-ncov) (accessed Nov 3, 2020).

4 World Health Organization. WHO Director-General’s opening remarks at the media briefing on COVID-19 - 11 March 2020. 2020; published online March 11. https://www.who.int/dg/speeches/detail/who-director-general-s-opening-remarks-at-the-media-briefing-on-covid-19---11-march-2020 (accessed July 24, 2020).

5 World Health Organization. WHO Coronavirus Disease (COVID-19) Dashboard. https://covid19.who.int (accessed Oct 16, 2020).

6 The World Bank. Population, total | Data. https://data.worldbank.org/indicator/SP.POP.TOTL (accessed July 24, 2020).

7 Wang D, Hu B, Hu C, et al. Clinical Characteristics of 138 Hospitalized Patients With 2019 Novel Coronavirus–Infected Pneumonia in Wuhan, China. JAMA 2020; 323: 1061–9.

8 Wan Y, Li J, Shen L, et al. Enteric involvement in hospitalised patients with COVID-19 outside Wuhan. Lancet Gastroenterol Hepatol 2020; 5: 534–5.

9 Xydakis MS, Dehgani-Mobaraki P, Holbrook EH, et al. Smell and taste dysfunction in patients with COVID-19. Lancet Infect Dis 2020; 20: 1015–6.

10 CDC. Evidence used to update the list of underlying medical conditions that increase a person’s risk of severe illness from COVID-19. Cent. Dis. Control Prev. 2020; published online Feb 11. https://www.cdc.gov/coronavirus/2019-ncov/need-extra-precautions/evidence-table.html (accessed Nov 4, 2020).

11 International Diabetes Federation. IDF Diabetes Atlas 9th edition 2019. 2019. https://www.diabetesatlas.org/en/ (accessed Nov 4, 2020).

12 Banik GR, Alqahtani AS, Booy R, Rashid H. Risk factors for severity and mortality in patients with MERS-CoV: Analysis of publicly available data from Saudi Arabia. Virol Sin 2016; 31: 81–4.

13 Yang JK, Feng Y, Yuan MY, et al. Plasma glucose levels and diabetes are independent predictors for mortality and morbidity in patients with SARS. Diabet Med J Br Diabet Assoc 2006; 23: 623–8.

14 Chee YJ, Tan SK, Yeoh E. Dissecting the Interaction between Coronavirus Disease 2019 and Diabetes Mellitus. J Diabetes Investig 2020; published online June 18. DOI:10.1111/jdi.13326.

15 Ministério da Saúde. SRAG 2020 - Banco de Dados de Síndrome Respiratória Aguda Grave - incluindo dados da COVID-19 - Open Data. https://opendatasus.saude.gov.br/dataset/bd-srag-2020 (accessed Nov 4, 2020).

16 Ministério da Saúde. Definição de Caso e Notificação. https://coronavirus.saude.gov.br/definicao-de-caso-e-notificacao (accessed Nov 4, 2020).

17 Ministério da Saúde. Guia de Vigilância Epidemiológica Emergência de Saúde Pública de Importância Nacional pela Doença pelo Coronavírus 2019. 2020;: 58.

18 Grant MC, Geoghegan L, Arbyn M, et al. The prevalence of symptoms in 24,410 adults infected by the novel coronavirus (SARS-CoV-2; COVID-19): A systematic review and meta-analysis of 148 studies from 9 countries. PLoS ONE 2020; 15. DOI:10.1371/journal.pone.0234765.

19 Yang J, Zheng Y, Gou X, et al. Prevalence of comorbidities and its effects in patients infected with SARS-CoV-2: a systematic review and meta-analysis. Int J Infect Dis 2020; 94: 91–5.

20 Di Domenico SL, Coen D, Bergamaschi M, et al. Clinical characteristics and respiratory support of 310 COVID-19 patients, diagnosed at the emergency room: a single-center retrospective study. Intern Emerg Med 2020; published online Nov 11. DOI:10.1007/s11739-020-02548-0.

21 Berenguer J, Ryan P, Rodríguez-Baño J, et al. Characteristics and predictors of death among 4035 consecutively hospitalized patients with COVID-19 in Spain. Clin Microbiol Infect 2020; 26: 1525–36.

22 Ghweil AA, Hassan MH, Khodeary A, et al. Characteristics, Outcomes and Indicators of Severity for COVID-19 Among Sample of ESNA Quarantine Hospital’s Patients, Egypt: A Retrospective Study. Infect Drug Resist 2020; 13: 2375–83.

23 Marson FAL. COVID-19 - 6 million cases worldwide and an overview of the diagnosis in Brazil: a tragedy to be announced. Diagn Microbiol Infect Dis 2020; 98: 115113.

24 The Lancet. COVID-19 in Brazil: “So what?” Lancet Lond Engl 2020; 395: 1461.

25 Lsy W, Exl L, Ayh K, Hx L, Pa T, Eh T. Age-Related Differences in Immunological Responses to SARS-CoV-2. J Allergy Clin Immunol Pract 2020; 8: 3251–8.

26 Rodríguez-Molinero A, Gálvez-Barrón C, Miñarro A, et al. Association between COVID-19 prognosis and disease presentation, comorbidities and chronic treatment of hospitalized patients. PLOS ONE 2020; 15: e0239571.

27 Ho FK, Petermann-Rocha F, Gray SR, et al. Is older age associated with COVID-19 mortality in the absence of other risk factors? General population cohort study of 470,034 participants. PLoS ONE 2020; 15. DOI:10.1371/journal.pone.0241824.

28 Garg S. Hospitalization Rates and Characteristics of Patients Hospitalized with Laboratory-Confirmed Coronavirus Disease 2019 — COVID-NET, 14 States, March 1–30, 2020. MMWR Morb Mortal Wkly Rep 2020; 69. DOI:10.15585/mmwr.mm6915e3.

29 Peckham H, de Gruijter NM, Raine C, et al. Male sex identified by global COVID-19 meta-analysis as a risk factor for death and ITU admission. Nat Commun 2020; 11: 6317.

30 Palaiodimos L, Kokkinidis DG, Li W, et al. Severe obesity, increasing age and male sex are independently associated with worse in-hospital outcomes, and higher in-hospital mortality, in a cohort of patients with COVID-19 in the Bronx, New York. Metabolism 2020; 108: 154262.

31 Hone T, Mirelman AJ, Rasella D, et al. Effect of economic recession and impact of health and social protection expenditures on adult mortality: a longitudinal analysis of 5565 Brazilian municipalities. 2019. DOI:10.1016/S2214-109X(19)30409-7.

32 Hone T, Rasella D, Barreto ML, Majeed A, Millett C. Association between expansion of primary healthcare and racial inequalities in mortality amenable to primary care in Brazil: A national longitudinal analysis. PLOS Med 2017; 14: e1002306.

33 de Oliveira BLCA, Luiz RR. Mortality by skin color/race and urbanity of Brazilian cities. Ethn Health 2017; 22: 372–88.

34 Baqui P, Bica I, Marra V, Ercole A, Schaar M van der. Ethnic and regional variations in hospital mortality from COVID-19 in Brazil: a cross-sectional observational study. Lancet Glob Health 2020; 8: e1018–26.

35 Zhang Y, Cui Y, Shen M, et al. Association of diabetes mellitus with disease severity and prognosis in COVID-19: A retrospective cohort study. Diabetes Res Clin Pract 2020; 165: 108227.

36 Pasquarelli-do-Nascimento G, Braz-de-Melo HA, Faria SS, Santos I de O, Kobinger GP, Magalhães KG. Hypercoagulopathy and Adipose Tissue Exacerbated Inflammation May Explain Higher Mortality in COVID-19 Patients With Obesity. Front Endocrinol 2020; 11. DOI:10.3389/fendo.2020.00530.

37 Ding Q, Shults NV, Harris BT, Suzuki YJ. Angiotensin-converting enzyme 2 (ACE2) is upregulated in Alzheimer’s disease brain. bioRxiv 2020;: 2020.10.08.331157.

38 Li M-Y, Li L, Zhang Y, Wang X-S. Expression of the SARS-CoV-2 cell receptor gene ACE2 in a wide variety of human tissues. Infect Dis Poverty 2020; 9: 45.

39 Bailey KL, Samuelson DR, Wyatt TA. Alcohol use disorder: A pre-existing condition for COVID-19? Alcohol Fayettev N 2021; 90: 11–7.

40 Zheng Z, Peng F, Xu B, et al. Risk factors of critical & mortal COVID-19 cases: A systematic literature review and meta-analysis. J Infect 2020; 81: e16–25.

41 Liang W, Guan W, Chen R, et al. Cancer patients in SARS-CoV-2 infection: a nationwide analysis in China. Lancet Oncol 2020; 21: 335–7.

42 Bae S, Kim SR, Kim M-N, Shim WJ, Park S-M. Impact of cardiovascular disease and risk factors on fatal outcomes in patients with COVID-19 according to age: a systematic review and meta-analysis. Heart 2020; published online Dec 16. DOI:10.1136/heartjnl-2020-317901.

